# Signatures of defective DNA repair and replication in early-onset renal cancer patients referred for germline genetic testing

**DOI:** 10.1101/2022.05.23.22275227

**Authors:** Elena V. Demidova, Ilya G. Serebriiskii, Ramilia Vlasenkova, Simon Kelow, Mark D. Andrake, Tiffiney R. Hartman, Tatiana Kent, Richard T. Pomerantz, Roland L. Dunbrack, Erica A. Golemis, Michael J. Hall, David Y.T. Chen, Mary B. Daly, Sanjeevani Arora

**Affiliations:** Cancer Prevention and Control Program, Fox Chase Cancer Center, Philadelphia PA, 19111; Kazan Federal University, Russia, 420008; Molecular Therapeutics Program, Fox Chase Cancer Center, Philadelphia PA, 19111; Department of Biochemistry and Molecular Biophysics, University of Pennsylvania, Philadelphia, PA,19104; Arcadia University, Glenside, PA; Department of Biochemistry & Molecular Biology, Sidney Kimmel Cancer Center, Thomas Jefferson University, Philadelphia, PA, 19107; Department of Cancer and Cellular Biology, Lewis Katz School of Medicine, Philadelphia, PA 19140; Department of Clinical Genetics, Fox Chase Cancer Center, Philadelphia PA, 19111; Department of Surgical Oncology, Fox Chase Cancer Center, Philadelphia PA, 19111; Department of Radiation Oncology, Fox Chase Cancer Center, Philadelphia PA, 19111

**Keywords:** early-onset cancer, renal cancer, DNA replication, DNA repair, germline

## Abstract

Early-onset renal cell carcinoma (eoRCC) is typically associated with pathogenic germline variants (PGVs) in RCC familial syndrome genes. However, most eoRCC patients lack PGVs in familial RCC genes and their genetic risk remains undefined. Here, we analyzed biospecimens from 22 eoRCC patients that were seen at our institution for genetic counseling and tested negative for PGVs in RCC familial syndrome genes. We performed whole-exome sequencing (WES) and found enrichment of candidate pathogenic germline variants in DNA repair and replication genes, including multiple DNA polymerases. Induction of DNA damage in peripheral blood monocytes (PBMCs) significantly elevated numbers of γH2AX foci, a marker of double-stranded breaks, in PBMCs from eoRCC patients versus PBMCs from matched cancer-free controls. Knockdown of candidate PGVs in Caki RCC cells increased γH2AX foci. Immortalized patient-derived B cells bearing candidate PGVs in DNA polymerase genes (*POLD1, POLH, POLE, POLK*) had DNA replication defects compared to control cells. Renal tumors carrying these DNA polymerase variants were microsatellite stable but had a high mutational burden. Direct biochemical analysis of the variant Pol δ and Pol η polymerases revealed defective enzymatic activities. Together, these results suggest that constitutional defects in DNA repair such as DNA replication repair underlie a subset of eoRCC cases. These findings may provide opportunities for use of the DNA repair targeting agents for eoRCC treatment. Screening patient lymphocytes to identify these defects may provide insight into mechanisms of carcinogenesis in a subset of genetically undefined eoRCCs.

**Significance Statement:** Screening for DNA repair variation may provide a more comprehensive risk assessment for eoRCC patients. Evaluation of DNA repair defects may also provide insight into the cancer initiation mechanisms for subsets of eoRCCs and lay the foundation for targeting DNA repair vulnerabilities in eoRCC.

## Introduction

Early onset renal cell carcinoma (eoRCC) in patients under the age of 60 has been increasing in frequency over the past decade (1). In the United States alone, the most recent analyses report a range of 3.0% annual increase in RCC incidence among individuals aged 45-49 years to as high as a 6.2% increase in incidence among those aged 25-29 years (1). EoRCC is in some cases linked to pathogenic germline variants (PGVs) in genes associated with RCC familial syndromes (*VHL, MET, FLCN, TSC1, TSC2, FH, SDHx, PTEN, BAP1*) (1-3); these genes are also often somatically mutated in sporadic RCC cases (2-5). Identification of a PGV in defined RCC familial syndrome genes guides clinical recommendations for surveillance, often improving survival due to early diagnosis of eoRCC. However, in recent work we found that only ∼3.7% of eoRCC patients undergoing cancer risk assessment report a PGV in the currently defined RCC familial syndrome genes (6), reflecting the fact that the majority of eoRCC cases remain genetically not well characterized. Currently, there are no National Comprehensive Cancer Network (NCCN) guidelines for detection, prevention, or risk reduction in individuals who present with an eoRCC but lack a PGV in a familial RCC gene (7).

Recently, we reported that a significant subset of eoRCC patients undergoing cancer risk assessment carry PGVs in DNA damage response and repair genes (∼8.55% vs. 3.7% in familial RCC genes) (6). Similarly, Carlo *et al*. reported an increased prevalence of PGVs in DNA repair genes in advanced clear cell and non-clear cell renal cancer patients (3, 8). Although PGVs in DNA repair genes are not currently defined by clinical testing guidelines as increasing risk of RCC, these recent studies suggest a potential role of defective DNA repair pathways in eoRCC carcinogenesis that could also lead to novel therapeutic options for RCC patients. Owing to the rising incidence of eoRCC and limited genetic data in younger RCC patients, we performed germline whole exome sequencing (WES) and functional assays on biospecimens from high-risk eoRCC patients diagnosed before 60 years of age, who were negative for PGVs in familial RCC syndrome genes and had a family history of RCC and/or other familial cancers. Our results suggest that constitutional defects in DNA repair, and specifically in function of DNA polymerases, underlie at least a subset of eoRCC cases. Screening patient lymphocytes to identify genotype-phenotype associations via functional assays may provide insight into the mechanism of carcinogenesis for a subset of genetically undiagnosed eoRCCs.

## Results

### eoRCC patients at the Fox Chase Cancer Center (FCCC) and family history of cancer

We analyzed the personal and family history of the probands in a cohort of 22 eoRCC patients. Multiple probands (6/22, 27%) had a second primary cancer, with breast cancer diagnosed in 3 probands (3/22, 14%) prior to diagnosis of RCC (**Table 1**). Here, 73% (n=16/22) of probands had a family history of RCC, with 50% (n=11/22) of probands having a first-degree relative with RCC. Intriguingly, 64% (n=14/22) of probands had a family history of cancers of the prostate, bladder, and thyroid, and melanoma, which have been associated with an RCC diagnosis (9).

**Table 1.** eoRCC patient characteristics, genomic findings and family history.

| Patient # | Sex | Genetic variants | Other cancers | Cancers in relatives |  |  |  |
| --- | --- | --- | --- | --- | --- | --- | --- |
|  |  |  |  | 1<sup>st</sup> degree | 2<sup>nd</sup> degree | 3<sup>rd</sup> degree | other |
| Pt 1 | M | *POLD1*<br>*POLH*<br>*MTOR*<br>*PARP1*<br>*FH*<br>*MCM2* | sarcoma, thyroid, Hodgkins lymphoma | SKIN, PANCREAS | COLON/RECTUM, THYROID, LUNG, PROSTATE, BREAST, BONE | SKIN, STOMACH |  |
| Pt 2 | F | *POLE*<br>*PDGFRA*<br>*BRCA2* |  | COLON | BREAST, COLON | RCC, BREAST |  |
| Pt 3 | F | *ATM*<br>*MITF* | breast | RCC, LUNG | BREAST, STOMACH |  | BLADDER, PANCREAS |
| Pt 4 | F | *RRM2B*<br>*BCL2L1* | breast | UTERUS | RCC, LEUKEMIA |  |  |
| Pt 5 | F | *OGG1*<br>*NEIL3*<br>*UBR5* | breast | SKIN |  | PROSTATE |  |
| Pt 6 | F | *RIF1*<br>*KDR*<br>*XRCC1* | neuroblast oma |  | MELANOMA, LUNG |  |  |
| Pt 7 | F | *MK167* | 2 primary RCC | COLON |  |  |  |
| Pt 8 | F | *RET*<br>*BCL2L1* |  | RCC |  |  |  |
| Pt 9 | F |  |  |  | RCC, BLADDER, TESTICLE/LIVER |  |  |
| Pt 10 | F | *PBRM1* |  | RCC, BREAST, PROSTATE |  |  | BREAST, COLON |
| Pt 11 | F |  |  | BREAST, STOMACH | BREAST, BLADDER, RCC |  |  |
| Pt 12 | M | *SCARB1* |  |  | RCC |  |  |
| Pt 13 | M | *TSC2* |  | LUNG, PANCREAS, SKIN |  |  |  |
| Pt 14 | M | *ATM (2)*<br>*FLT3*<br>*SMARCA4* |  | RCC, BLADDER, LUNG |  |  |  |
| Pt 15 | M | *EGF* |  | RCC, BLADDER, LIVER, PROSTATE |  |  |  |
| Pt 16 | M | *POLK*<br>*EXO1*<br>*BRCA2*<br>*MUTYH* |  | BLADDER |  |  | RCC |
| Pt 17 | M |  |  | RCC |  |  |  |
| Pt 18 | M | *NFX1* |  | RCC, PROSTATE |  |  |  |
| Pt 19 | M | *SDHB*<br>*NDUFA13*<br>*MMP9* |  | RCC, BLADDER, COL.POLYPS | BLADDER |  |  |
| Pt 20 | M | *MMP9*<br>*MSH3*<br>*LTK*<br>*POLR2A* |  | RCC, LUNG, NON-HODGKINS LYMPH |  |  |  |
| Pt 21 | M |  |  | BREAST, RCC |  |  |  |
| Pt 22 | M | *FLT4*<br>*SMARCE1* |  | RCC | LUNG |  |  |

F – female, M – male, Pt – patient, RCC – renal cell carcinoma

### Analysis of whole-exome sequencing data reveals enrichment of germline variation in DNA repair and replication genes in eoRCC patients

We performed WES on lymphocyte DNA from the 22 eoRCC probands, detecting candidate PGVs based on a candidate gene list that was developed in our prior studies ((10); see (**Supplementary Table 1**) and Supplementary Materials and Methods). Briefly, the candidate gene list was developed by a comprehensive hypothesis-driven framework with the following assumptions: 1) genes involved in genome stability (using Gene Ontology terms such as DNA repair, DNA replication, DNA damage checkpoints, cell cycle, mitotic machinery, replication stress, DNA damage response, chromatin remodeling) would be important for hereditary cancer risk (5, 11-13); and 2) an expanded network of genes relevant to renal biology (such as cellular metabolism) and genes somatically mutated in RCC that might be relevant for eoRCC-predisposition (5, 11, 13).

Novel *candidate* PGVs were stringently defined as those that are predicted to disrupt protein function by the consensus of at least 4 protein predictor algorithms; are rare (gnomAD allele frequency<0.01); and are nonsynonymous variants (frameshifts, stop gains, and splicing) (see Supplementary Methods for variant prioritization). We identified candidate PGVs in 18/22 eoRCC patients in the study, yielding a total of 44 variants in 39 genes (**Table 1, and Supplementary Table 2**). Gene Ontology analysis confirmed that the candidate PGVs were enriched in DNA repair and replication pathway genes (**Figure 1A and Supplementary Figure 1**, WebGestalt) (14). Here, 10 patients (46%; 10/22) had 17 candidate PGVs in 14 genes currently associated with hereditary cancers across major organ systems (*ATM, BRCA2, POLD1, POLE, FH, MITF, MSH3, MUTYH, PDGFRA, RET, SDHB, SMARCA4, SMARCE1, TSC2*). Only 4 patients had candidate PGVs in RCC familial syndrome genes (18%, 4/22 – *FH, MITF, SDHB, TSC2*). Finally, a total of 14 patients (64%; 14/22) had candidate PGVs from our expanded candidate gene list, from genes not currently defined as RCC-predisposing.

**Figure 1.**
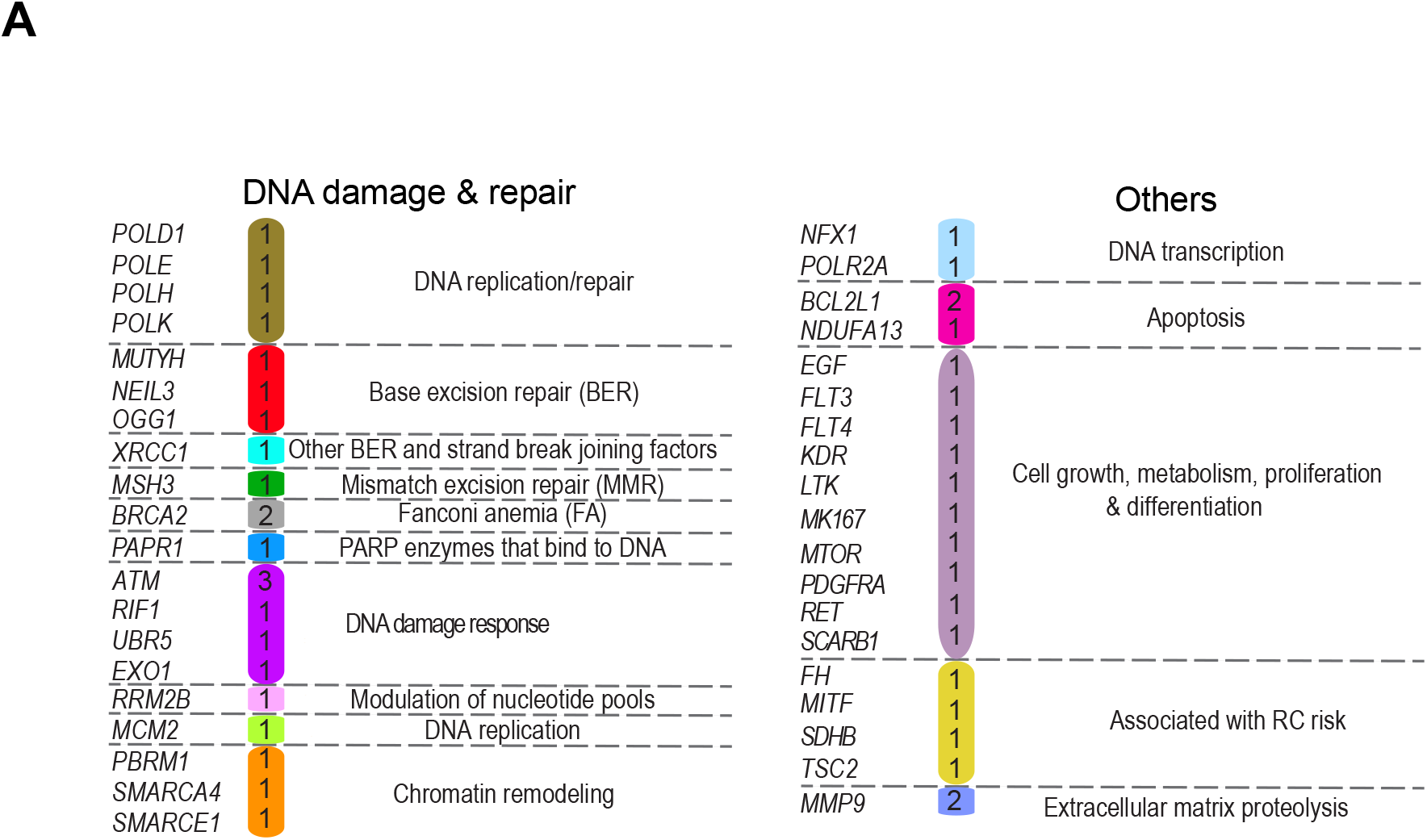
Enrichment of predicted pathogenic variants in DNA repair genes in the cohort. **A**. Summary of variants in genes and pathways, identified in the cohort. In color - number of variants identified for each gene. For detailed information, see Supplementary tables 1 and 2.

Among the DNA repair-associated genes, candidate PGVs were found in *BRCA2* (2/22 patients; 9%, **Table 1, Supplementary Table 2**) and in *ATM* (2/22 patients; 9%, **Table 1, Supplementary Table 2**). In addition, 5 candidate variants in DNA replication-repair genes (4/22 patients; 18%, **Table 1, Supplementary Table 2**, *POLD1* and *POLH* (Pt #1), *POLE* (Pt #2), *POLK* (Pt #16), and *RRM2B* (Pt #4)). Pt #1 had candidate missense variants in PolD1, a catalytic subunit of the replicative DNA polymerase, Pol δ, and in the translesion synthesis DNA polymerase, Pol η. *POLD1* G2275A p.V759I is in a highly conserved region of PolD1 subunit of the Pol δ (coded by *POLD1* gene) (15) and occurs at a high allele frequency in the Ashkenazi Jewish population (0.0213) reported in gnomAD (versus 0.0018 in the complete gnomAD dataset (16)). *POLH* G626T (p.G209V) is in the Pol η catalytic core (17, 18). Pt #2 has a candidate stop-gain G4872A (p.W1624X) variant in the *POLE* gene, coding replicative DNA polymerase Pol ε, in the conserved C-terminal domain (19). Pt #4 had a splice site (intronic) variant in *RRM2B*, coding a subunit of p53-inducible ribonucleotide reductase, which performs *de novo* conversion of ribonucleotide diphosphates into the corresponding deoxyribonucleotide diphosphates for DNA synthesis (20), in genome position #103237248. Pt#16 has a missense variant in the highly conserved N-terminus domain of another translesion synthesis polymerase, *POLK* G85A (p.E29K); this variant has been described previously as compromising enzyme activity (21). ClinVar classifications of the variants are presented in **Supplementary Table 2**, with most variants currently classified as variants of uncertain significance (VUS).

### Primary lymphocytes from eoRCC patients have reduced capacity to suppress DNA double strand breaks (DSBs)

To begin to assess the functional effect of candidate PGVs in genes linked to DNA replication and repair, we assessed the numbers of γ(phospho)-H2AX foci (a marker of DSBs, (22)) in patient peripheral blood monocytes (PBMCs) at baseline and after treatment with the DNA polymerase inhibitor aphidicolin (**Figure 2A**). In PBMCs from both matched cancer-free controls (by age and gender) and eoRCC patients, aphidicolin significantly elevated the number of γH2AX foci; however, aphidicolin-treated cells from eoRCC patients had markedly higher numbers of γH2AX foci than those from similarly treated controls on treatment, indicating reduced DSB repair mechanism in eoRCC patient cells (**Figure 2A**, P<0.001). In complementary work, we tested whether the genes bearing candidate PGVs were specifically needed to suppress DNA DSBs in RCC cells. For this, we used siRNA to deplete the *POLD1, POLE, POLH, POLK, RRM2B*, and *ATM* genes in the Caki RCC cell line. For each gene, knockdown significantly increased γH2AX foci relative to control (**Supplementary Figure 2**) further supporting a role for these proteins in DSB repair in renal cells.

**Figure 2.**
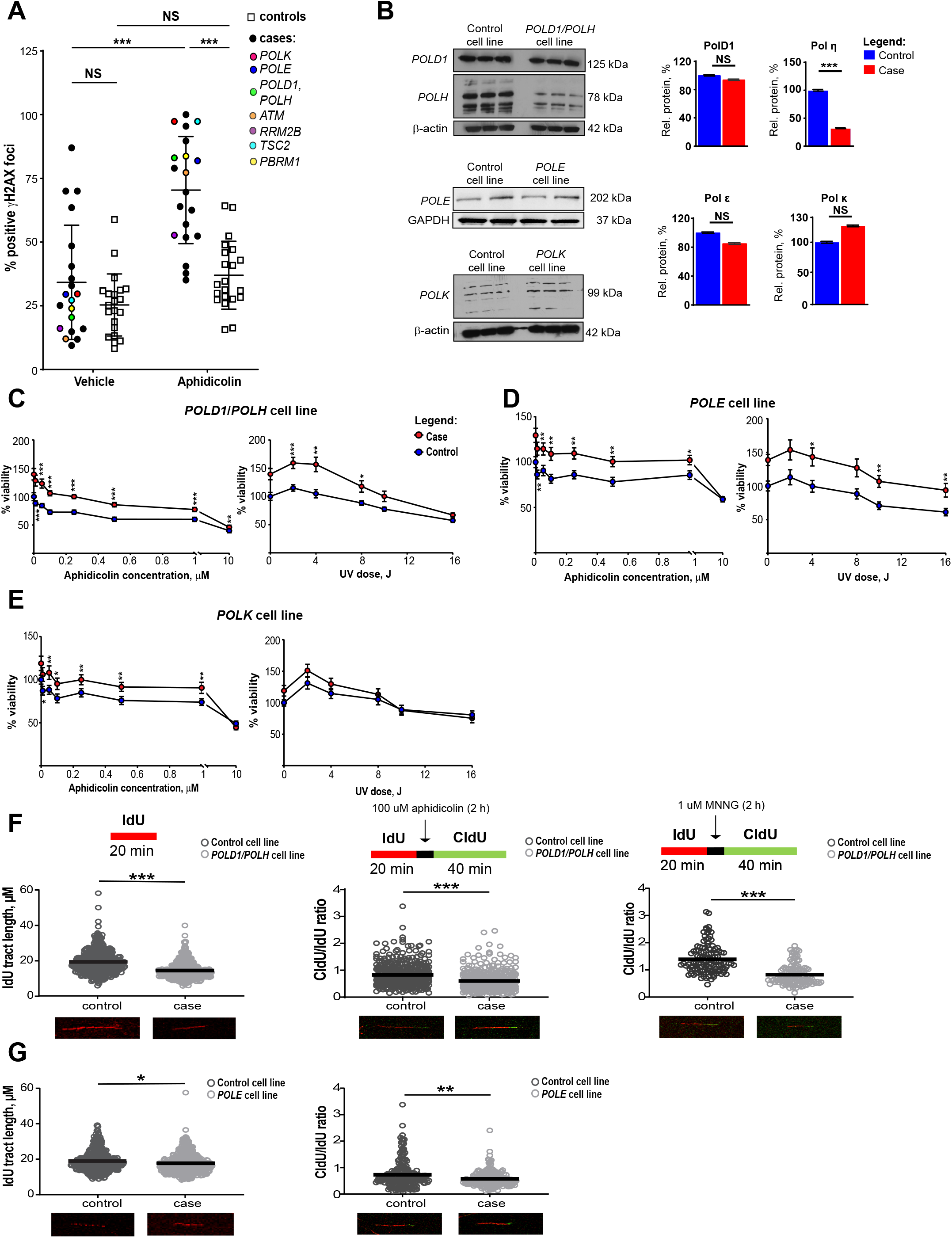
Cell-based functional analysis revealed defects in DNA repair and DNA replication in lymphocytes from eoRCC patients. **A**. γH2AX foci immune fluorescence staining in primary PBMCs from eoRCC patients versus matched controls, at baseline or post treatment with aphidicolin (2h). PBMCs from patients showed statistically significant elevation of γH2AX foci post treatment with aphidicolin. Data were normalized and are presented as percent of positive γH2AX foci. **B**. Representative Western blots showing expression of PolD1, Pol η, Pol ε and Pol κ in EBV-transformed cell lines carrying variants versus matched controls (without the variants). Data quantification was performed based on 3 independent biological repeats, technical repeats are presented on gels. **C-E**. Relative viability of EBV-transformed cell lines was assessed by CTB assay at baseline or after treatment with aphidicolin or UV. Data were normalized to CTB values for controls and are presented as percent cellular viability for *POLD1*/*POLH* (**C**), *POLE* (**D**), and *POLK* (**E**) cell lines. Data from 3 independent biological repeats are presented. **F-G**. Difference in DNA replication fork elongation/restoration in EBV-transformed cell lines (**F** - *POLD1, POLH*; **G** - *POLE* lines) at the baseline and post replications stress was assessed using DNA fiber assay. At baseline the EBV-transformed cells were labeled with IdU for 20 min, for fork restoration cells then were treated with 100 μM aphidicolin or 1 uM MNNG for 2 h, and then labeled with CldU for 40 min. For all conditions, post labeling, cells were lysed, and DNA fibers stretched onto glass-slides, fixed, denatured, blocked, and stained with corresponding antibodies. Fiber images were captured using the Nikon TS2R Inverted Microscope and analyzed in ImageJ software. Data for 3 independent repeats are presented as IdU tract length or CldU/IdU tract length ratio. For all graphs: *** for p<0.001, ** for p<0.01, * for p<0.05 and NS for p>0.05, unpaired, non-parametric t-test, Mann-Whitney criteria.

### Patient-derived cell lines with candidate PGVs in DNA polymerases have DNA replication defects

We prepared EBV-transformed cell lines from the primary lymphocytes of 3 patients bearing candidate PGVs (henceforth referred to as the *POLD1/POLH* cell line, *POLE* cell line, and *POLK* cell line) and from several age- and gender-matched cancer-free controls. The *POLD1/POLH* cell line had significantly reduced levels of the Pol η; for the other candidate PGVs, polymerase level was not affected (**Figure 2B)**. Cell Titer Blue (CTB) cellular assays showed significantly better metabolic capacity or cellular viability than control-derived cell lines when treated either with aphidicolin, or with ultraviolet light (which causes bulky adducts in DNA), suggesting that cell lines from patients had better ability to tolerate DNA damage (**Figures 2C-E**). Analysis of cell cycle did not show any significant differences in patients and matched control cell lines (**Supplementary Figure 3A**).

**Figure 3.**
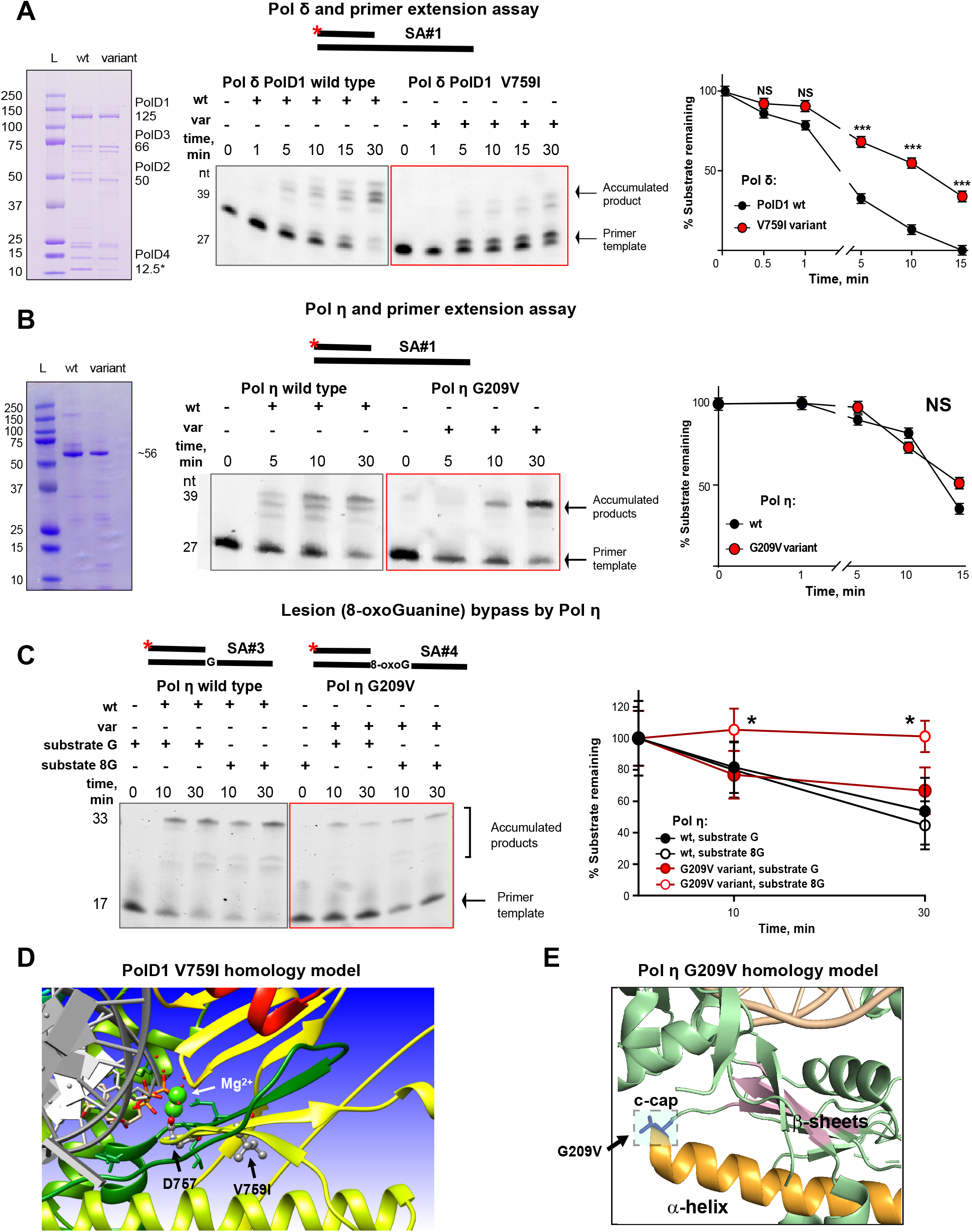
Structural and biochemical assays revealed altered enzymatic activities of the PolD1 and Pol η variants. **A**. Pol δ complex and primer extension assay. On the left - representative gel image of purified wt and variant Pol δ protein complexes, containing 4 subunits: PolD1 (125 kDa), PolD2 (50 kDa), PolD3 (66 kDa) and PolD4 (12.5 kDa). Center and right – Pol δ complex primer extension assay with quantification. Representative gel image showing reactions performed with 20 nM Cy-3 labeled DNA-duplex template (SA#1), 20 nM of indicated proteins and 500 uM dNTPs. PolD1 V759I complex extended DNA-template less efficiently comparing to wt protein complex. Data for 3 independent repeats are presented. **B**. Pol η and primer extension assay. On the left - representative gel image of purified wt and variant Pol η catalytic cores (432 amino acids), molecular weight ∼56 kDa. Center and right – Pol η primer extension assay with quantification. Representative gel image showing reactions performed with 20 nM Cy-3 labeled DNA-duplex template (SA#1), 20 nM of indicated proteins and 500 uM dNTPs. Data for 3 independent repeats are presented. **C**. Pol η lesion (8-oxoG) bypass assay with quantification. Representative gel image showing reactions performed with 20 nM Cy-3 labeled DNA-duplex template with 8-oxoG in the position, opposite to 3‵-OH group (SA#4), and template of the same sequence without lesion (SA#3), 20 nM of indicated proteins and 500 uM dNTPs. Data for 3 independent repeats are presented. For **A-C**: *** for p<0.001, ** for p<0.01, * for p<0.05 and NS for p>0.05, unpaired, non-parametric t-test, Mann-Whitney criteria. All template sequences may be found in **Supplementary Table 4. D-E**. Homology modeling structures using yeast protein templates for human PolD1 V759I (**D**) and for human Pol η G209V (**E**).

To further expand on these cell-based findings, we used a DNA fiber assay (see Supplementary Methods) and directly compared DNA replication in patient-derived versus control cells, either untreated or following treatment with aphidicolin or with a DNA-alkylating agent, 1-Methyl-3-nitro-1-nitrosoguanidine (MNNG) (**Figure 2F-G Supplementary Figure 4A-D)**. The *POLD1*/*POLH* cell line and the *POLE* cell line exhibited a significantly lower rate of DNA replication in untreated cells (∼1.4-fold decrease, p<0.001 for the *POLD1*/*POLH* cell line, and ∼1.9-fold decrease, p<0.05 for the *POLE* cell line versus controls). We also observed a significantly lower replication fork recovery after 2h treatment with aphidicolin (∼1.44-fold decrease, p<0.001 for the *POLD1*/*POLH* cell line, and ∼1.88-fold decrease, p<0.01 for the *POLE* cell line versus controls) (**Figure 2G-H)**. Intriguingly, the *POLD1*/*POLH* cell line showed defective replication fork restoration (∼1.2-fold decrease, p<0.001 versus control line) 2h post-treatment with MNNG (**Figure 2G**). The *POLK* cell line did not show any defects in DNA replication and replication recovery under the conditions tested (**Supplementary Figure 4A)**. A complete summary of results for the DNA polymerase variants is provided in **Supplementary Table 3**.

**Figure 4.**
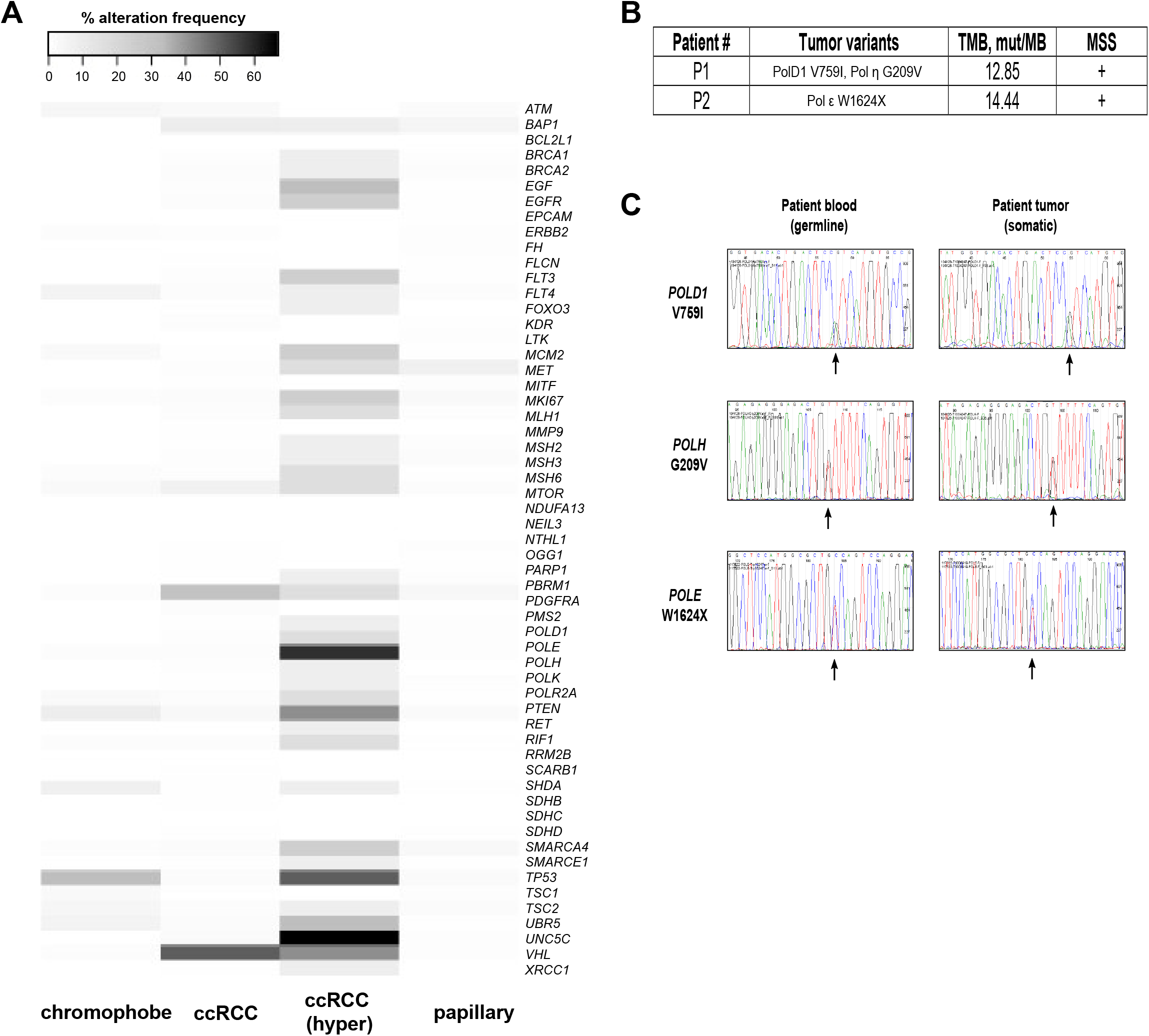
Renal tumors carrying polymerase variants showed high TMB, MSS, and no LOH. **A**. Percent alteration frequency in 897 tumors from TCGA in different histological types of RCC: chromophobe (n=66), ccRCC - clear cell renal cell carcinoma (n=538), ccRCC (hyper) - hypermutated samples (n=12), papillary (n=293). **B**. TMB and MSS data are presented for Pt #1 (*POLD1* V759I, *POLH* G209V) and Pt #2 (*POLE* W1624X). **C**. Tumor and normal Sanger sequencing for variants in Pt #1 (*POLD1* V759I, *POLH* G209V) and Pt #2 (*POLE* W1624X) showing no LOH. Arrows show variants of interest on sequencing tracks.

### Altered enzymatic activity of Pol δ and Pol η variant proteins

Among the candidate PGVs detected in polymerases, the Pol κ variant E29K has previously been biochemically shown to possess not only a significantly reduced catalytic efficiency but also reduced replication fidelity (21). E29K is in a conserved region of the Pol κ N-terminus, the N-clasp subdomain (1-32 aa), which is essential to maintaining the stability of the open conformation of the Pol κ active site (23). Intriguingly, a previous study showed that deletion of the first 67 amino acids reduces Pol κ activity during translesion synthesis (TLS, i.e., replication by efficient bypass of bulky lesions in DNA) (24).

To directly test effects of the other candidate PGVs on polymerase activity, we first purified the polymerase delta (Pol δ) protein complex, with the PolD1 (*POLD1*), PolD2 (*POLD2*), PolD3 (*POLD3*), and PolD4 (*POLD4*) subunits from recombinant protein co-expressed in *E. coli*, and with preparations containing either wild type (wt) PolD1 or PolD1 V759I variant (**Figure 3**). Both the wt and the variant-containing Pol δ complexes extended a Cy3-labeled DNA primer-template; however, the V579I variant complex had significantly less robust polymerase activity than the wt complex (**Figure 3A**, p<0.001). Furthermore, when Pol δ complexes containing PolD1 wt or PolD1 V759I proteins were mixed in a ratio of 1:1, the appearance of the extended primer-template was significantly more robust than the variant alone but significantly less robust than the wt alone. This result suggests that the variant is not only impaired for function but has a partial dominance over the wt in this assay (**Supplementary Figure 5**, p<0.001).

**Figure 5.**
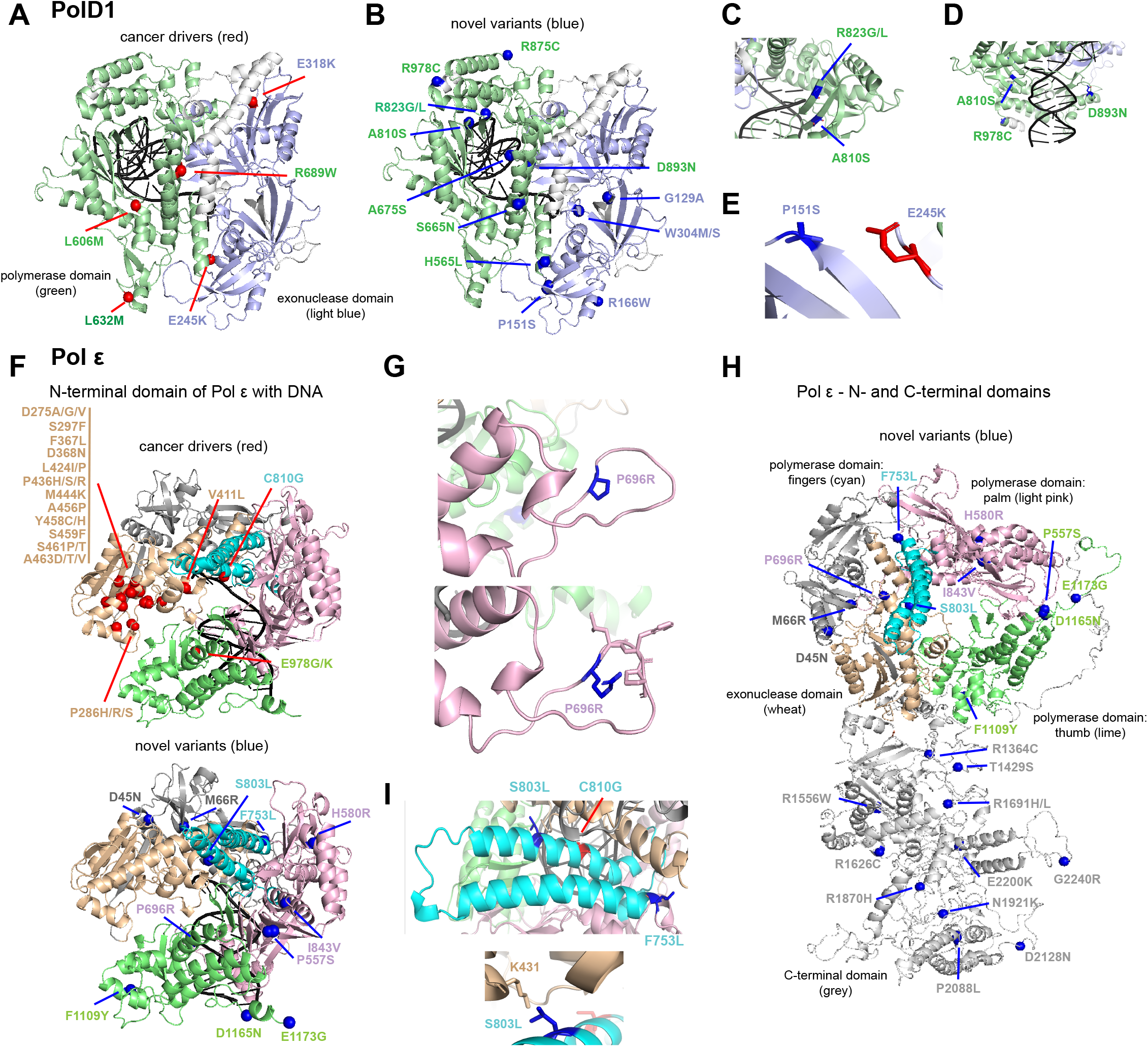
Structure mapping of the novel PolD1 and Pol ε variants from hypermutated ccRCCs in TCGA. **A-E**. DNA-bound PolD1 3D-model was refined from PDB:3IAY. The colored functional domains are exonuclease (light blue, residues 131-477) and polymerase (green, residues 550-978). **A**. Red spheres represent known cancer drivers. **B**. Blue spheres represent variants of uncertain significance in ccRCC. **C-E**. Fragments of PolD1 model showing variants: A810S and R823G (**C**), D893N, A810S, and R978C (**D**), P151S and E245K (**E**). **F**. DNA-bound N-terminal domain of Pol ε was refined from PDB:4M8O. The colored functional domains are N-terminal subdomain (dark grey, residues 31-281), exonuclease (wheat, residues 282-527), polymerase (light pink, palm: 528-950; cyan, fingers: 769-833; lime, thumb: 951-1186). Red spheres represent known cancer drivers (structure above). Blue spheres represent variants of uncertain significance in ccRCC (structure below). **G**. A fragment of Pol ε model showing variant P696L. **H**. 3D-model of whole-length Pol ε (without DNA). Structure was refined as described in Methods based on (26). The colored functional domains are N-terminal subdomain (dark grey, residues 31-281), exonuclease (wheat, residues 282-527), polymerase (light pink, palm: 528-950; cyan, fingers: 769-833; lime, thumb: 951-1186), C-terminal domain (light grey, residues 1308-2222). Blue spheres represent variants of uncertain significance in ccRCC. **I**. Fragments of Pol ε model showing variants S803L and F753.

Pol η is a low fidelity polymerase, which contributes to its ability to perform TLS (25). Hence, G209V variant and wt Pol η prepared in *E. coli* were assessed for their ability to extend labeled DNA primer-template duplexes (**Figures 3B-C**). In the absence of DNA damage (e.g., in a normally base-paired template), the wt and variant proteins both extended the template (**Figure 3B**, p>0.05), but the observed bands suggest higher processivity for the variant on template without lesions compared to wt (**Figure 3B**). To evaluate repair of DNA damage, TLS activity was also tested using a template containing an 8-oxoGuanine (8-oxoG) DNA lesion (25). Pol η wt bypassed the 8-oxoG lesion robustly compared to the Pol η variant (p<0.05, **Figure 3C**), suggesting better processivity for the wt protein on template with DNA lesion (25).

Finally, biochemical analysis of a purified Pol ε variant, W1624X, was not performed as it is a stopgain variant in the C-terminal domain or CTD, truncating 662 amino acids of the protein. The CTD region is not well-studied, but is thought to be essential for stability of the Pol ε holoenzyme (26).

### Structural modeling of DNA polymerase variants in eoRCC suggests impact on polymerase function

The PolD1 V759I variant is targets a residue two amino acids away from residue D757 (**Figure 3D**, in dark green). In the PolD1 active site, D757 coordinates the Mg^2+^ ions (neon green spheres) required for DNA synthesis and plays a direct role in the catalytic mechanism and binding of DNA (15). Structural modeling indicates that a substitution of the valine (V) 759 to isoleucine (I) could plausibly alter the position of D757 and disrupt the efficiency of DNA polymerization. To further understand the structural changes that each polymerase variant might induce, we calculated the change in stability of each amino acid substitution as described in Supplemental Methods and **Supplementary Table 4**. Interestingly, the PolD1 I759 yielded a mild stabilization (**ΔΔG** of -1.36 kcal/mole) relative to the wildtype V759. The I759 residue makes twice the number of hydrophobic contacts as the wildtype V759 with the long helix below. The variant could lock this strand in an overly rigid position that compromises DNA polymerization steps that involve flexibility, consistent with the biochemical results observed in **Figure 3A**.

The Pol η G209 residue is in the catalytic core of the polymerase (**Figure 3E**, residues 1-432, colored green) at a position often called the C-cap, i.e. the residue in this position is proximal to the C-terminal end of the α-helix (in orange) (27). Typically, valine, threonine, and isoleucine residues are not preferred in the C-cap, due to poor solvation at the C-terminus of the helix when the side chains are bulky (27). Structural modeling of the G209V substitution showed that the valine with a bulkier side chain could not only alter the stability of the α-helix, but also the nearby β-strands of the catalytic active site (in pink). Rosetta modeling shows the variant G209V is capable of making 5 hydrophobic contacts across the cleft with R24, which is just downstream from the key residue D13, which coordinates active site metals that are central to the catalytic mechanism and the binding of the incoming NTP. The predicted change in stability of the Pol η G209V variant revealed a significant destabilization (**ΔΔG** of 5.67 kcal/mole) relative to the wildtype G209, consistent with lower levels observed in protein purification (**Figure 2B, Figure 3B** (left gel), and **Supplementary Table 4**). Interestingly, the activity of the G209V compares well with wt for a normal primer template and might even be more processive (**Figure 3B**, middle gel). However, the variant appears defective for TLS when the template contains an 8oxoG (**Figure 3C**). Consideration of these data together show that changes in stability and conformation may be subtle and even result in an alteration of substrate preference. Thus, a careful combination of both computational and biochemical methods is required to gain a clear understanding of the role a given polymerase variant might have in DNA replication, repair, and cancer initiation and/or progression.

### EoRCCs carrying candidate PGVs in DNA polymerases are hypermutant and microsatellite stable (MSS)

To extend these functional tests, we next explored tumor mutation burden (TMB) in tumors from RCC patients in TCGA and from the FCCC eoRCC patients. Previous studies have shown that colorectal and endometrial tumors carrying mutations in *POLE* exonuclease domain (ExoD) and in *POLD1* exhibit a high burden of mutations, are typically MSS but few cases with microsatellite instability (MSI) have been reported, and do not exhibit loss of heterozygosity (LOH) (15, 28-36). RCCs are typically non-hypermutated, with an average TMB of ∼1 mut/Mb (37); however, rare hypermutated (≥10 mut/Mb) and rarer ultra-hypermutated (>100 mut/Mb) RCCs carrying polymerase mutations, with or without MSI, have been reported (37). Analysis of renal tumors in TCGA found that several candidate PGV genes from this study (**Figure 4A**), including specifically DNA polymerase genes were mutated in these hypermutant clear cell RCCs (ccRCCs) s (**Figure 4A**). We analyzed the TMB and MSI/MSS status of tumors from the FCCC eoRCC patients and found hypermutation without MSI (*POLD1/POLH* tumor with 12.85 mut/Mb; *POLE* tumor 14.44 mut/Mb) (**Figure 4B**). The tumors from the FCCC eoRCC patients did not exhibit LOH of the polymerase genes, as has been observed in other polymerase-mutated tumors (37, 38) (**Figure 4C**).

To expand the analysis of DNA polymerases in RCC, we next modeled the structural consequences of the PolD1 and Pol ε variants in hypermutated ccRCCs from TCGA. **Supplementary Table 5** shows the predicted changes in stability for 20 different PolD1 variants and 23 Pol ε variants from hypermutated ccRCCs in TCGA. A broad range of both stabilizing and destabilizing variants was found in all domains of each of these polymerases. **Figure 5** shows these variants in relation to known pathogenic variants in *POLD1* (**Figure 5A-E**) and *POLE* (**Figure 5F-I**). PolD1 R823G/L is in a β-sheet of the polymerase domain close to the DNA (**Figure 5C**). The substitution of a positively charged arginine (R) to a hydrophobic glycine (G) or leucine (L) could destabilize the β-sheet and impact DNA binding. PolD1 D893N is positioned close to the DNA and a variant in this region may destabilize DNA binding or position for DNA-protein interactions (**Figure 5D**). PolD1 P151S is in a β-sheet in the ExoD and the change from proline (P) to serine (S) could destabilize the β-sheet geometry (**Figure 5E**). P151 is close to a known cancer driver mutation (37, 39), E245K, in the unfolded region of the ExoD (**Figure 5A**).

Pol ε P696R is in the palm region of the polymerase domain, which is highly conserved among replicative polymerases (**Figure 5G)**. Arginine (R) has a very large positively charged side chain when compared to smaller proline (P), suggesting this variant may disrupt the polymerase structure and impact DNA synthesis. Pol ε S803L and F753L are in the flexible region of the polymerase domain or the fingers (in cyan, **Figure 5I**). This finger region shifts (27° tilt) on DNA binding (40), and thus plays an essential role in polymerase function. S803 is close to the positively charged lysine (K) 431 in the ExoD, and serine (S) is polar and a smaller residue than the hydrophobic leucine (L). Pol ε S803L is near a site of known cancer driving mutations, C810 (37), suggesting that specific alterations in this α-helix could impact polymerase functioning. F753L is on the border of the fingers and the polymerase domain, close to the ExoD and could be important in the coordination of these regions with or without DNA binding. Finally, several variants were found in the C-terminal domain (**Figure 5H**, in light grey), which is currently not well-studied, but is known for stabilizing the Pol2 (human Pol ε) complex in yeast (26).

## Discussion

In this study, we focus on analysis of candidate PGVs in DNA repair and replication genes in probands with RCC diagnosed prior to 60 years of age undergoing cancer risk assessment at our cancer center who tested negative for RCC familial syndrome genes. We applied a well-curated pipeline of candidate genes in genome stability, metabolism, metabolic stress, normal renal function, RCC biology, and chromatin remodeling to germline WES data from eoRCC patients. Gene Ontology analysis confirmed that the identified candidate PGVs were enriched in DNA repair and replication genes. Intriguingly, we found that many eoRCC patients exhibit defects in suppression of DSBs in their primary PBMCs, with PBMCs from eoRCC patients exhibiting higher γH2AX foci than matched cancer-free controls in response to DNA damage. Direct knockdown of some of these candidate variant genes in Caki RCC cell line also led to increased γH2AX foci. Genes with candidate PGVs were found to be mutated in sporadic RCCs, with specific enrichment of alterations in *BRCA2*, and in DNA polymerase (*POLE, POLD1, POLH1*, and *POLK*) genes in hypermutant RCCs in TCGA. Importantly, detailed analysis of candidate PGVs in DNA polymerase genes from the FCCC eoRCC patients confirmed the damaging nature of the candidate PGVs and suggested a mechanistic basis for association of these variants with observed defects in DNA repair and replication. Several PolD1 and Pol ε variants from the hypermutant RCCs in TCGA were proximal to the catalytic center or substrate binding regions and will benefit from similar future biochemistry experiments shown in this study.

This work complements a number of recent studies indicating inherited defects in DNA replication machinery may increase cancer risk. PGVs in the ExoD of *POLE* and *POLD1* predispose to cancer and exhibit a strong mutagenic effect, however, the role of non-ExoD variants in cancer risk has been controversial (29, 31, 41). A recent study reported that the *POLD1* candidate PGV (p.V759I, in Pt #1) is frequently present in the Ashkenazi Jewish population and proposed this gene as a founder mutation (2). Mertz et. al. have demonstrated a strong mutator effect of the PolD1 polymerase domain variant R689W in human cells (42). Several TLS polymerases including Pol η and Pol κ, are important for preventing accumulation of single strand DNA gaps, and the replication of DNA fragile sites. Owing to their high error-propensity, TLS polymerases are likely to contribute to oncogene-induced mutagenesis (43, 44).

It is likely that the candidate PGVs in DNA replication and repair genes detected here interact with other germline variants to impact eoRCC risk. In this study, Pt #1 also harbored candidate PGVs in *MTOR, PARP1, FH, MCM2*, suggesting the *POLD1* and *POLH* variants assessed here may act together with other variants to augment the DNA repair defects observed. Pt#2 with a *POLE* candidate PGV also harbored candidate PGVs in *BRCA2, PDGFRA*. In other studies, hypermutant ccRCCs also carried mutations in *TP53, PTEN, VHL*, and *UNC5C* (45, 46). It is also possible that defects in polymerase genes affect cancer risk by affecting biological processes beyond DNA replication and repair. We found that the immortalized cell lines from Pt#1, 2, and 16 (*POLK*) showed better metabolic capacity compared to control cell lines. Intriguingly, patients with mandibular hypoplasia, deafness and progeroid features with concomitant lipodystrophy, characterized by germline PGVs in *POLD1*, also present with mitochondrial dysfunction and metabolic abnormalities (15, 47, 48). Conversely, some familial RCC genes (such as *FH, VHL, PBMR1*, and *SDHx*) have also been implicated in suppression of DSBs and in replication stress (49-52), based on mechanisms that are not well understood.

It is important to note that while the family history of the high-risk probands in this study is suggestive of underlying genetics (53), the clinical testing using a RCC familial syndrome genes or panel did not yield any actionable PGVs according to current NCCN recommended guidelines. Here, 64% of probands had an extensive family history of cancers of the prostate, bladder, and thyroid, and melanoma, all of which have previously been associated with RCC diagnosis (9). In fact, PGVs in DNA repair genes have been reported as risk factors for bladder, skin, thyroid, and prostate cancers (54-59). Our results suggest that in the absence of PGVs in RCC familial syndrome genes or phenotypic features of familial RCC or family history of RCC, a comprehensive assessment of general cancer predisposition genes, including DNA repair genes, may be beneficial. Here, multiple probands (27%) had at least one additional primary cancer with breast cancer being the most common additional primary cancer (14%). A recent retrospective analysis of the Swedish Cancer Registry showed that ∼10% of RCC patients develop another second primary cancer, and this is currently thought to be independent of the primary RCC, suggesting broader cancer predisposition (60), compatible with PGVs in genes affecting DNA repair.

Besides genetic screening, these data suggest the value of functional assessment for the families of individuals with eoRCC. In this study, the majority of eoRCC PBMC biospecimens samples exhibited elevated γH2AX levels and candidate variants in DNA repair genes. Our data support a potential role of germline variation in DNA repair/replication leading to suboptimal encoding protein activity, and genome instability. Overall, these data suggest that assays of γH2AX foci in normal cells, supporting germline variation in DNA repair/replication genes, could be a potential tool for the identification of individuals with genetically unexplained eoRCC. As these defects could be detected in normal cells, it could lead to the identification of individuals in need of cancer risk assessment. This is especially relevant because case-control studies suggest that an elevated familial RCC risk may be multifactorial, and or due to an interaction of the heritable genetics and the shared environment (53). It is possible that defective DNA repair in the heterozygous state could be a recessive heritable factor that when combined with other RCC risk factors may jointly increase the risk of eoRCC.

Currently, the therapeutic significance of DNA repair genes is not clinically defined for RCC. Evidence is emerging that PARP inhibitors could be therapeutics of choice in RCCs that may not carry mutations in the classical *BRCA* genes, but which have other defects in DNA repair, with recent clinical trials assessing the use of PARP inhibitors in RCC (61, 62). Hence, there is a critical need to not only understand the biological impact of defective DNA repair in renal tissue but to also define risk of RCC due to a germline defect in DNA repair genes. Further exploration in a larger and more diverse (by race/ethnicity) patient population is clearly of interest for future work. In sum, study of PBMCs offers a useful approach to resolve the biological and clinical significance of rare gene variants identified by exome sequencing and may improve clinical approaches to risk assessment and medical management.

## Materials and Methods

### eoRCC patient population, and peripheral blood DNA analysis

Case-only eoRCC probands that underwent clinical germline genetic testing between 2010-2016 were included in this study (n=22). Patients were followed by the Genitourinary Program at the Fox Chase Cancer Center and had undergone evaluation for inherited cancer risk at the FCCC Family Risk Assessment Program (RAP). Each participant had a strong family cancer history as shown in **Table 1**, with either multiple first-degree or second-degree relatives with RC, RC-associated cancers, or other cancers. The mean age at eoRCC diagnosis was 48 years (range 36–59 years). No pathogenic mutations were identified from sequencing the following RC-specific genes: *VHL, MET, FLCN, TSC1, TSC2, FH, SDHx, PTEN* and *BAP1*. The patients had consented to the FCCC RAP Registry, which allowed further research, genomic sequencing, and banking of their biospecimens in the BioSample Repository Facility. The patients reported here were self-reported white, non-Hispanic. Family histories were obtained by trained licensed genetic counselors and verified by attending physicians. The analyses performed and publication of deidentified information was under the approval of the FCCC Institutional Review Board Committee protocol number 14-831. See **Supplementary Methods** for more methods.

## Data Availability

All data produced in the present work are contained in the manuscript. Only the DNA sequence reads data is not available for sharing as consent was not obtained for sharing.

## Abbreviations

8-oxoG,: 8-oxoGuanine;
BWA,: Burrows-Wheeler aligner;
ccRCC,: clear cell renal cell carcinoma;
CTB,: Cell Titer Blue;
CldU,: 5-Chloro-2’-deoxyuridine;
DSBs,: double-strand breaks;
eoRCC,: early-onset renal cell carcinoma;
DDR,: DNA damage response;
EBV,: Epstein-Barr virus;
ExAC,: Exome Aggregation Consortium;
ExoD,: exonuclease domain;
FCCC,: Fox Chase Cancer Center;
FDR,: false discovery rate;
GATK,: genome analysis toolkit;
GnomAD,: Genome Aggregation Database;
GO,: gene ontology;
IdU,: 5-Iodo-2′-deoxyuridine;
LOH,: loss of heterozygosity;
MMR,: mismatch repair;
MNNG,: 1-Methyl-3-nitro-1-nitrosoguanidine;
MSI,: microsatellite instability;
MSS,: microsatellite stability;
NCCN,: National Comprehensive Cancer Network;
PBMCs,: peripheral blood monocytes;
PGVs,: pathogenic germline variants;
Pt,: patient;
RAP,: risk assessment program;
RCC,: renal cell carcinoma;
SNPs,: single nucleotide polymorphisms;
TLS,: translesion synthesis;
TMB,: tumor mutation burden;
VUS,: variant of uncertain significance;
WebGestalt,: WEB-based GEne SeT AnaLysis Toolkit;
WES,: whole-exome sequencing;
wt,: wild type.

## Acknowledgments

The work in this grant was supported by the resources and expertise provided by the FCCC Cell Culture, Biosample Repository, Genomics, Biostatistics, Population Sciences, Molecular Modelling, and High Throughput Screening Facilities. We acknowledge Nina Shah (summer undergraduate at FCCC) for assistance with knockdown experiments, Emmanuelle Nicolas (Genomics Facility, FCCC) and Waleed Iqbal (Drexel University, PA) for assistance with genomic analysis. We express gratitude to Lisa Bealin, Andrea F. Forman, and Kim Rainey (Department of Clinical Genetics, FCCC) for valuable assistance on the study.

## Grant Support

All Fox Chase Cancer Center affiliated authors are in part supported by the NCI Core Grant, P30 CA006927, to the Fox Chase Cancer Center. S.A. was supported by the DOD W81XWH-18-1-0148, and a CEP Award from the Yale Head and Neck Cancer SPORE. M.J.H. was supported by funding from the American Cancer Society. M.B.D was supported by the NIH U01 CA164920, R01 CA207365 grants. E.A.G. was supported by NIH R01 DK108195. E.V.D. and I.G.S. were partially supported by the Kazan Federal University Strategic Academic Leadership Program (PRIORITY-2030). R.J.D. was supported by NIH R35 GM122517. R.T.P. was supported by NIH grants 1R01GM130889 and 1R01GM137124.

## Notes

**Disclosures/Conflict of Interest**. M.J.H. performs collaborative research (with no funding) with the following: Myriad Genetics, Invitae Corporation, Ambry Genetics, Foundation Medicine, Inc. He also performs collaborative research (with no funding) and is part of a Precision Oncology Alliance funded by Caris Life Sciences (cover travel and meals at meetings). S.A. performs collaborative research (with no funding) with Caris Life Sciences, Foundation Medicine, Inc., Ambry Genetics, and Invitae Corporation. S.A.’s spouse is employed by Akoya Biosciences and has stocks in Akoya Biosciences, HTG Molecular Diagnostics, Abcam Plc., and Senzo Health. S.A., M.J.H., E.A.G., I.G.S. have patents and/or pending patents related to cancer diagnostics/treatment. All other authors declare no competing interests.

### Competing Interest Statement

M.J.H. performs collaborative research (with no funding) with the following: Myriad Genetics, Invitae Corporation, Ambry Genetics, Foundation Medicine Inc. He also performs collaborative research (with no funding) and is part of a Precision Oncology Alliance funded by Caris Life Sciences (cover travel and meals at meetings). S.A. performs collaborative research (with no funding) with Caris Life Sciences, Foundation Medicine, Inc., Ambry Genetics, and Invitae Corporation. S.A. spouse is employed by Akoya Biosciences and has stocks in Akoya Biosciences, HTG Molecular Diagnostics, Abcam Plc., and Senzo Health. S.A., M.J.H., E.A.G., I.G.S. have patents and/or pending patents related to cancer diagnostics/treatment. All other authors declare no competing interests.

### Author Declarations

Ethics committee/IRB of Fox Chase Cancer Center gave ethical approval for this work.

