## Supplemental Text for "Signatures of defective DNA repair and replication in early-onset renal cancer patients referred for germline genetic testing"

### Supplementary text.

#### Supplementary Methods.

#### Gene variants annotation and prioritization

PBMC DNA from the 22 eoRCC patients was sent to BGI Americas Corporation (Cambridge, MA) for WES at 100X average coverage. Agilent V5 exome capture kit was used for exon capture (Agilent Technologies, Wilmington, DE). Library preparations were done using Illumina standard protocol. Each captured library was indexed, then loaded onto Hiseq4000 platform (Illumina, CA) for 100 bp paired-end high-throughput sequencing. Sequence reads were mapped to human reference genome (hg19) using the Burrows-Wheeler Aligner (BWA)(1). Single Nucleotide Polymorphisms (SNPs) and small Insertion/Deletions (InDels) were detected using Genome Analysis Toolkit (GATK)(2). Variant annotation, filtering and prioritization was performed as previously described(3). Briefly, variant annotations were computed with ANNOVAR(4) version 2019-10-24 and include: population frequency, from ANNOVAR-provided versions of the gnomAD genome and exome call sets(5) predicted protein impact, using ANNOVAR-provided versions of RefSeq(6) predicted deleteriousness, using the ANNOVAR file dbnsfp30a(7).

As previously described(3), selected annotations were applied to filter and prioritize the variants prior to manual evaluation. (a) Hard-filtering step to exclude variants highly unlikely to be causative. Variants were required to have genotype quality ≥10, read depth ≥10, and maximum population frequency <0.01. (b) Variants were required to be predicted to impact the protein sequence derived from any RefSeq transcript, or to be previously reported as pathogenic or likely pathogenic by at least one of the variant effect prediction tools (SIFT, POLYPHEN2, LRT, MutationTaster, MutationAssessor, or CADD) or reported in ClinVar(3, 8). (c) All candidate variants were extremely rare as determined by examination of representation in the Genome Aggregation Database (gnomAD)(9). (d) As previously described(3), for a candidate gene approach, datasets of RCC-related genes obtained from multiple sources, both commercial and publicly available were integrated. RCC-related genes were obtained from Ingenuity(10), TCGA (using cBioPortal(11-14)), DisGeNet(15), ICGC(16), HGMD(17), GeneCards(18), and OMIM(19) Professional databases; genes implicated in multiple cancers (cancer census genes) were obtained from COSMIC database(13) and our previous publication(1, 2). The candidate gene list was developed by a comprehensive hypothesis-driven framework with the following assumptions: 1) genes involved in genome stability (using Gene Ontology terms such as DNA repair, DNA replication, DNA damage checkpoints, cell cycle, mitotic machinery, replication stress, DNA damage response, chromatin remodeling) would be important for general hereditary cancer risk(20-23), and 2) an expanded network of genes relevant to renal biology (such as cellular metabolism) and renal cancer(20, 22, 23). Subsequently, the gene candidates were prioritized by building a network using Cytoscape program and MiMi plugin, and subsequently only retaining the genes that were either nominated by two or more sources or nominated by one database but interacting with than two or more high-confidence genes. **Supplementary Table 1** summarizes the genes in the candidate gene list. All prioritized gene variants were confirmed by examining the exome reads for sequence quality and variant representation in gnomAD(9) and/or dbSNP, and variants of interest by direct Sanger sequencing. WebGestalt tool was used for gene set enrichment analysis to extract biological insights from the genes of interest(24). The online WebGestalt tool was used, and an over representation analysis was performed. The enrichment results were prioritized by significant *p*-values, and FDR thresholds at 0.01. Finally, MSI or MSS (from clinical PCR-based testing and/or IHC-testing for mismatch repair proteins) was extracted from the patient's clinical data.

**Analysis of RCC studies in TCGA**

cBioPortal (http://www.cbioportal.org) was used to access data for the candidate genes (*ATM, BCL2L1, BRCA1, BRCA2, EGF, EGFR, ERBB2, FH, FLT3, FLT4, FOXO3, KDR, LTK, MCM2, MKI67, MMP9, MSH3, MTOR, NDUFA13, NEIL3, NTHL1, NXF1, OGG1, PARP1, PBRM1, PDGFRA, POLD1, POLE, POLH, POLK, POLR2A, RET, RIF1, RRM2B, SCARB1, SDHB, SMARCA4, SMARCE1, TSC2, UBR5, UNC5C, XRCC1, BAP1, FH, FLCN, MET, MITF, PTEN, SDHA, SDHB, SDHC, SDHD, TSC1, TSC2, VHL, TP53, MLH1, MSH2, PMS2, EPCAM, MSH6*) from the most recent TCGA studies (data downloaded in December 2019). The studies used were Kidney Chromophobe (n = 66), Kidney Renal Clear Cell Carcinoma (n = 538), Kidney Renal Papillary Cell Carcinoma (n = 293). All the studies were TCGA Firehouse Legacy datasets. For the analysis, we calculated the TMB in each sample. Mutation counts were adjusted by dividing each result by candidate gene length. These samples were analyzed separately. Data has been processed using RStudio and GraphPad Prism 9.

**Modelling of DNA polymerase variant interactions**

The structural information on Pol ε family of proteins comes mainly from yeast structures found in the PDB, obtained by both X-ray diffraction and cryo-EM methods. The X-ray diffraction structure of the N-terminal lobe in the presence of DNA (PDB code: 4M8O)(25) provided high resolution data for the preliminary model of the human protein discussed here (using Swiss-Model(26)). The more recent cryo-EM structure of the entire yeast Pol2 (analogue for human Pol ε) in complex with the other subunits (PDB code: 6WJV)(27) provided a lower resolution template for modeling the whole human Pol ε with a view to portions that interact with the other subunits of the holoenzyme. There was also a structure of the human Pol ε C-terminal residues 2142 to 2286 (PDB code: 5VBN)(28) that is in complex with the B-subunit of the holoenzyme. To refine our homology models, we captured the best structural data available for each portion of Pol ε and generated two models of the human protein in the presence and absence of DNA. Model refinement methods Rosetta Relax and SCWRL4(29, 30) were used to optimize side chain rotamer interactions with other subunits, substrate DNA and incoming NTP's. UCSF Chimera(31) was used to align models and published structures, to assess residue contacts and hydrogen bonding, and to rationalize mechanistic effects of novel variants on a variety of protein functions, including interactions with other subunits in the holoenzyme.

DNA-bound polymerase structures for PolD1 and Pol η structure motifs was generated by I-TASSER(32-34), then refined in PYMOL (http://www.pymol.org) and then further refined based on the results of the SWISS-model(26) and by using the most homologous structures in the PDB. For PolD1, *Saccharomyces cerevisiae* (PDB code: 3IAY) structure and for Pol η *Saccharomyces cerevisiae* (PDB code: 4O3P) was used. For PolD1 the refined structure shown is from amino acid residues 96-985. The colored functional motifs are exonuclease (residues 130-477) and the polymerase domain (residues 550-978). Protein domain regions were based on pfam (http://pfam.xfam.org/family/PF00136).

Known cancer drivers and novel variants in PolD1 and Pol ε were mapped on to the respective homology models. Pol ε whole-length model was used to represent novel variants, found in C-terminal domain, and shorter model was used to represent location of variants when binding DNA. Novel Pol ε and PolD1 variants were obtained from kidney cancer studies in TCGA (http://www.cbioportal.org).

**Predicting the change in stability of missense variants**

In order to assess the effects of missense variants on protein stability, we used the Rosetta Molecular Modeling Suite module named "ddg-monomer"(35) (meaning ddG of the variant relative to the wild type amino acid). For this module to be a reliable method of predicting stability changes, the input protein structure model should be complete with no chain breaks. As the initial models built from various model organism templates contain gaps and incomplete loops, we chose to utilize AlphaFold2 models for each of the Pol ε, PolD1, and Pol η. The company DeepMind has made structure predictions of the entire human proteome freely available from an EMBL-EBI server (https://alphafold.ebi.ac.uk/). Prior to using the ddg-monomer module, each of the alphafold models were minimized with required constraints by another Rosetta minimization tool.

The purpose of the ddg-monomer module is to predict the change in stability (the ddG) of a monomeric protein induced by a point mutation. The application takes as input the pre-minimized wild-type model and generates a structural model of the point-mutant. The ddG is given by the difference in Rosetta energy between the wild-type structure and the point mutant structure. The application was run using the high-resolution protocol (using the current Rosetta Scoring Function [beta_nov15] and for specific settings see the application file in **Supplementary Table 7**), and 25 models each of the wild-type and mutant structures were generated, and the most accurate ddG is taken as the difference between the mean of the top-3-scoring wild type structures and the top-3-scoring point-mutant structures. Rosetta follows the convention that negative ddG values indicate increased stability i.e., ddG = mutant energy - wildtype energy.

The ddg-monomer application was used to predict stability changes for the PolD1 V759I variant, Pol η G209V variant, 20 different PolD1 variants, and 23 Pol ε variants from TCGA (see **Supplementary Tables 4 and 5**). The change in stability of the variant relative to the wildtype residue is listed in the ddG column, and the color coding is from green (stabilizing) to white to red (destabilizing). It should be noted that the Alphafold models are apoprotein structures and the Rosetta ddg-monomer calculations are performed without consideration of DNA or metal ligands. While there are minor shifts in the position of a few secondary structure elements, the change in stabilities calculated on the apoprotein models are equivalent to variants in the homology model with DNA (For example when comparing the Alphafold PolD1 apoprotein model to a homology model built in the presence of DNA substrate, the RMSD between 586 alpha carbon atom pairs is 1.16 square angstroms).

**Protein expression and purification**

Polymerase delta complex expression vectors were obtained from Dr. Pomerantz at the Thomas Jefferson University in Philadelphia. Vectors information could be found in(36). Pol η expression plasmid were purchased from GenScript (Piscataway, NJ), and contains the cDNA sequence corresponding to 1-711 amino acid inserted into a pET32a backbone. QuikChange Lightning Multi Site-Directed Mutagenesis Kit (Agilent Technologies, CA) was used for site-directed mutagenesis, changes were confirmed by sequencing performed by Genewiz (South Plainfield, NJ). BL21-CodonPlus(DE3)-RIL competent cells (Agilent Technologies, CA) were used as expression strain. Pol δ wild type (wt) and Pol δ containing the PolD1 V759I variant protein complexes were expressed and purified as described(36) before with minor changes. Briefly, cell lysis was performed by 10 cycles with microfluidizer at the 10-15 psi pressure. Pol η wt and Pol η G209V variant were expressed and purified as described(37, 38) with minor change: instead of MonoS column and buffer C, dialysis was performed before concentration with 2 L of the following buffer (25 mM sodium phosphate (pH 7.4), 10% glycerol, 200 mM NaCl, 5 mM β-mercaptoethanol). Identified protein fractions were analyzed by SDS-PAGE gels. Selected fractions containing highly purified polymerase were aliquoted and frozen and stored at −80 °C until further use.

**Primer extension, single nucleotide incorporation and 8-oxoG DNA lesion bypass.**

The reactions were performed at 37°C with 20 nM Cy3/5-labeled oligonucleotide substrates (see **Supplementary Table 6)**, 25 mM Tris-HCl (pH 8.8), 1 mM DTT, 0.01% Igepal, 0.1 mg/ml BSA, 10% glycerol, 10 mM $MgCl_2$, 20 nM of enzyme (Pol δ wild type complex or Pol δ PolD1 V759I complex, Pol η wild type or Pol η G209V), and 500 µM dNTPs(37, 39, 40). Reactions were terminated by adding 10 µl of stop solution (45 mM Tris-HCl, 45 nM Boric acid, 1 mM EDTA, 6% Ficoll Type 400, 3,5 M Urea, 0.005% Xylene Cyanol). The products were separated by electrophoresis in a 15% denaturing polyacrylamide gel (Invitrogen, MA), and detected by PharosX Plus Imager (BioRAD, CA) and quantified using ImageJ and GraphPad Prism 8 software. All substrates were purchased as duplexes from Integrated DNA Technologies (Coralville, IA).

**Assays with primary PBMCs culture, patient- or control-derived EBV cell lines.**

PBMCs were collected from patients and controls (cancer-free, age and gender-matched) in identical fashion and preserved by standard methods as previously described(1). B-cells from patients and normal controls were immortalized with EBV as previously described(1, 41). Briefly, lymphocytes isolated from 10 ml blood by centrifugation over Ficoll-Paque were resuspended in 2.5 ml complete RPMI-1640 without insulin. Immortalization was initiated by the addition of EBV strain B95-8 as 1 ml of filtered supernatant from the marmoset B-cell line GM 7404. After incubation for 2 h at 37°C, 6.5 ml of complete RPMI containing 1 mg/ml cyclosporine A (Sigma-Aldrich, MO) was added and the suspension cultured at 37°C, 5% CO2 in an up-right flask. After 3 weeks, the culture was split, and 5 ml fresh medium added to each flask. After an additional week of incubation, cells in one of the flask pairs were cryopreserved and cells in the second flask were subcultured for eventual harvest. The assays performed using either the primary PBMCs or EBV-transformed patient or control derived cell lines are described below.

DDR assays were performed using the primary PBMCs as previously described(1, 2). Primary PBMCs were available from 20 of the exome-sequenced patients and 20 age-matched and gender-matched individuals without a cancer diagnosis or a family history of cancer. Control samples were obtained from the FCCC Biosample Repository Facility. Briefly, cells were cultured in RPMI-1640 containing 15% fetal bovine serum (HyClone Laboratories, Logan, UT), 2 mM L-glutamine (Life Technologies, Grand Island, NY), 50 µM 2-mercaptoethanol (Sigma-Aldrich, St. Louis, MO), 0.2 units human recombinant insulin (Sigma-Aldrich, St. Louis, MO) per ml, 50 units penicillin and 50 mg streptomycin per ml (complete RPMI), and then stimulated with phytohemagglutinin (PHA)-M (Life Technologies, Grand Island, NY) and recombinant human interleukin 2 (IL-2) (NCI Preclinical Repository) for 72 h. For immunofluorescence, cells were allowed to attach to poly-d-lysine-coated 96-well plates, and then treated with vehicle, or 20 µM aphidicolin, and fixed in paraformaldehyde 2 hours later, permeabilized, blocked and then stained with anti-γH2AX antibody (#05–636, Millipore, Temecula, CA). After the primary antibody, cells were stained with the secondary anti-mouse antibody (Cell Signaling Technology Inc., Danvers, MA) and counter stained with DAPI. Cells were imaged on the 6 ImageXpress Micro automated microscope (Molecular Devices, Sunnyvale, CA) and analyzed by MetaXpress software. Foci were scored for γH2AX staining, and the results were displayed and exported using the AcuityXpress software package (Molecular Devices, Sunnyvale, CA).

Protein expression studies were performed using EBV-immortalized cell lines from patient and matched controls. Briefly, whole cell lysates were prepared, and Western blot analysis was performed for the respective proteins and β-actin or GAPDH loading control. Total protein levels were quantified and normalized to the loading control. Primary antibodies used were: PolD1 (ab186407, Abcam, Cambridge, MA), Pol ε (#MA5-13616**,** Thermo Fisher Scientific, Waltham, MA), Pol η (#13848, Cell Signaling Technology Inc., Danvers, MA), Pol κ (A301-975A-T, Santa Cruz Biotechnology Inc., CA), β-actin (ab20272, Abcam, Cambridge, MA), GAPDH (from Loading Control Ab sampler kit, #5142T, Cell Signaling Technology Inc., Danvers, MA).

For cell cycle analysis, immortalized EBV cells were fixed in 96% ethanol at $1x10^6$ cells. Then cells were washed in PBS and finally stained with PI/RNAse Staining buffer (cat. #550825, BD Biosciences, NJ). The Guava PCA (Millipore, IL) for cell cycle analysis.

For siRNA studies, Caki cells were plated in 96-well plates and then transfected with GL2 (negative control), WRN (positive control) or two independent siRNAs (5nM) for each protein. Cells were then fixed in paraformaldehyde, permeabilized, blocked and then stained with anti-γH2AX antibody (#05–636, Millipore, Temecula, CA). After the primary antibody, cells were stained with the secondary anti-mouse antibody (Cell Signaling Technology Inc., Danvers, MA) and counter stained with DAPI. Cells were imaged on the 6 ImageXpress Micro automated microscope (Molecular Devices, Sunnyvale, CA) and analyzed by MetaXpress software. Foci were scored for γH2AX staining, and the results were displayed and exported using the

AcuityXpress software package (Molecular Devices, Sunnyvale, CA). The data are plotted as relative induction of γH2AX to GL2 control from 2 independent experiments.

Cellular viability experiments were performed using the EBV lines from patient and matched controls. Here, cells were plated in 96-well plates and were treated for 2h with vehicle (cell culture medium) or with aphidicolin (Sigma-Aldrich, St. Louis, MO), or Ultraviolet (UV) light at concentrations or doses shown respectively. At 72h post-treatment, CellTiterBlue (#G8080, Promega Corporation, Madison, WI) was added and the absorbance was read using a Perkin Elmer Plate Reader (PerkinElmer Inc, Waltham, MA). The signal was read using a Perkin Elmer Plate Reader.

DNA fiber assay was performed as previously described(42, 43). Briefly, patient-derived EBV cell lines carrying original variants in DNA polymerase (Pt #1 - PolD1 V759I/Pol η G209V, Pt #2 – Pol ε W1624X, P t#16 – Pol κ E29K, see **Table 1**) were labelled with 250 µM of 5-Iodo-2′-deoxyuridine or IdU (Sigma-Aldrich, MO) or/and 50 µM of 5-Chloro-2'-deoxyuridine or CldU based on experimental conditions (see **Figure 2** and **Supplementary Figure 4**). Post labeling, the cell lines were pelleted, re-suspended in ice cold PBS buffer (~3000 cells/µl), then 2.5 µl cells (~ 9000 cells) were spotted onto a glass slide (Superfrost Plus, ThermoFisher Scientific, Waltham, MA), and mixed with 7.5 µl of lysis buffer (200 mM Tris-HCl pH7.4, 50 mM EDTA, 0.5% SDS). For DNA fibers, cells were lysed for 8 min and then the slides were tilted at 45° angle and air-dried. DNA fibers on the slide were fixed with a methanol: acetic acid (3:1) solution for 10 min, and then slides were rinsed with water, air-dried and frozen overnight. Next day, the DNA fiber slides were washed with PBS, DNA was denatured using 2.5 M HCl for 2.5 h, washed in PBST (PBS & 0.1% Tween-20), and then blocked with 2% BSA for 40 min at RT. After blocking, the fibers slides were incubated with anti-IdU/anti-CldU antibody (1:100, cat. # 347580, BD Biosciences, NJ & 1:500, cat. #NB500-169, Novus Biologicals, CO, correspondingly) followed by a secondary antibody (1:300, cat. #A-11062 anti-mouse Alexa594 & cat. #A-11006 anti-rat Alexa488, ThermoFisher Scientific, Waltham, MA). The slides were mounted using ProLong Diamond Antifade Mountant (cat. # P36970, ThermoFisher Scientific, Waltham, MA) and imaged using the Leica SP8 (Leica Microsystems Inc., Buffalo Grove, IL) and Nikon TS2R Inverted Microscope (Nikon Instruments Inc., Melville, NY) microscopes, and the data were analyzed using the ImageJ & GraphPad Prism 9 software.

**Tumor DNA analysis**

DNA extraction from FFPE tumor tissue was performed by the FCCC BioSample Repository Facility following standard protocols and as previously described(3, 44). Briefly, tumor cells were collected from an H&E-stained section that was demarcated by a pathologist. The cells were deparaffinized by incubation with xylene at 56°C for 1 hr. Xylene was removed and the cells were washed with descending concentrations of ethanol and the pellet was allowed to dry at 56°C for 10 min. This was followed by DNA isolation using the QIAmp DNA micro kit (Qiagen catalog number 56304). The isolated DNA was measured on Nanodrop and used for further experiments. Sanger sequencing was also performed using tumor or lymphocyte DNA, where appropriate, for multiple candidate variants (**Supplementary Table 2**). DNA was sent to BGI Americas Corporation (Cambridge, MA) for WES using Agilent V6 exome kit (Agilent Technologies, Wilmington, DE). The resulting sequence data were filtered for any low-quality reads, aligned to the human reference genome (UCSC H19 build), and assessed for sequencing quality. SNPs and Indels were called, annotated, and validated as per the standard WES bioinformatic pipeline. BGI also calculated the Tumor Mutation Burden or TMB as the total number of somatic mutations in the WES data. To obtain an accurate measure of TMB, all non-tumor and benign common mutations were filtered out based on the normal PBMC WES data from each patient and against well-annotated databases such as 1000 Genomes Project, dbSNP, and COSMIC, as is standard practice(45, 46). To report mutations per megabase of DNA, 38 Mb was used as the estimate of the exome size and nonsynonymous mutations were used to report TMB, as previously described(46). Tumors with TMB > 10 mut/Mb were considered hypermutated(47).

#### Statistics

Where appropriate we used unpaired or paired non-parametric t-tests, Mann-Whitney, or Wilcoxon criteria for the assessment of the differences between cases and controls in yH2AX immunofluorescent staining, protein expression, cell counts, cell cycle analysis, CTB assay, DNA fiber assay and biochemical assays. P-value less 0.05 was considered significant and significant findings are represented as *** for $p<0.001$, ** for $p<0.01$, * for $p<0.05$ and NS for $p>0.05$.

#### Supplementary References.

**Supplementary Figure Legends.**

**Supplementary Figure 1. Pathway enrichment for the genes with identified DDR germline variants according to over-representation analysis (ORA).** FDR is provided inside the boxes of the heat map.

**Supplementary Figure 2. A. siRNA depletion of *POLD1, POLE, POLH, POLK, RRM2B* and *ATM* genes in Caki RCC cell line.** A. Cells were transfected with the designated siRNAs (two per gene), or GL2 control or *WRN* positive control. Cells were fixed, permeabilized, blocked and stained for γH2AX antibody. Cells were scored for γH2AX foci and the data are plotted as relative induction of γH2AX to GL2 control from 2 independent experiments. *** for $p<0.001$, ** for $p<0.01$, * for $p<0.05$ and NS for $p>0.05$, Wilcoxon signed rank test.

**Supplementary Figure 3. Representative cell cycle data from immortalized B-cell lines from Pt #1 (*POLD1*, *POLH*), Pt#2 (*POLE*), Pt#16 (*POLK*) PBMCs and corresponding immortalized B-cell lines from matched control. A.** Percent positive gated cells is presented for each cell cycle phase for each cell line (controls on the left in blue, cases on the right in red). Data for 3 independent repeats are presented. *** for $p<0.001$, ** for $p<0.01$, * for $p<0.05$, NS for $p>0.05$, Wilcoxon signed rank test.

**Supplementary Figure 4. Difference in DNA replication fork elongation/restoration in EBV-transformed cell line with *POLK* (Pt #16) variant, and representative pictures of DNA fibers for the main figures 2E and 2F.** For all graphs: *** for $p<0.001$, ** for $p<0.01$, * for $p<0.05$ and NS for $p>0.05$, unpaired, non-parametric t-test, Mann-Whitney criteria. Means with SD are plotted. Data for 3 independent repeats are presented as IdU tract length or CldU/IdU tract length ratio. **A.** Difference in DNA replication fork elongation/restoration in EBV-transformed cell line with *POLK* E29K variant at the baseline and replication stress was assessed using DNA fiber assay. At baseline the EBV-transformed cells were labeled with IdU for 20 min, for fork restoration cells then were treated with 100 µM aphidicolin and then labeled with CldU for 40 minFor all conditions, after labeling, cells were lysed, and DNA fibers stretched onto glass-slides, fixed, denatured, blocked, and stained with corresponding antibodies. Fiber images were captured using the Nikon TS2R Inverted Microscope and analyzed in ImageJ software. **B-D.** Additional representative pictures for each DNA fiber experiment: **B** for IdU 20 min labeling (main figures **2E**, **2F,** current figure **A);** **C** for short aphidicolin treatment (main figures **2E, 2F,** current figure **A**); **D** for treatment with MNNG (main figure **2E**). Scale bar = 5 µm.

**Supplementary Figure 5. Pol δ complex primer extension competition assay with quantification.** Representative gel image showing reactions performed with 20 nM Cy-3 labeled DNA-duplex template (SA#1), 20 nM of indicated proteins (wild type complex, V759I complex and both wild type + V759 variant complexes in ratio 1:1) and 500 uM dNTPs. Under presence of the both wild type and V759I enzymes, DNA-template was extended less efficiently compared to wild type and more efficiently compared to V759I complexes alone. Data for 3 independent repeats are presented. *** for $p<0.001$, ** for $p<0.01$, * for $p<0.05$ and NS for $p>0.05$, unpaired, non-parametric t-test, Mann-Whitney criteria.

**Supplemental Table Legends.**

**Supplementary table 1.** List of candidate genes for WES analysis.

**Supplementary table 2.** Annotation of candidate variants identified in the 22 eoRCC patients.

**Supplementary table 3**. Complete summary of results for the DNA polymerase variants identified in the eoRCC patients.

**Supplementary table 4.** Protein stability or ddG values for the PolD1 V759I and Pol η G209V variant proteins that were assessed biochemically in the study.

**Supplementary table 5.** *POLE* and *POLD1* variants in hypermutated ccRCC in TCGA. The table shows the variants from TCGA with allele counts typically observed in the GnomAD database, TMBs observed in association with these variants in other studies, and finally protein stability or ddG values for the variant proteins.

**Supplementary table 6.** DNA substrates used in biochemical assays.

**Supplementary table 7.** ddG application file.
