## Supplementary figures and images for "Signatures of defective DNA repair and replication in early-onset renal cancer patients referred for germline genetic testing"

### supp fig 1

**A**

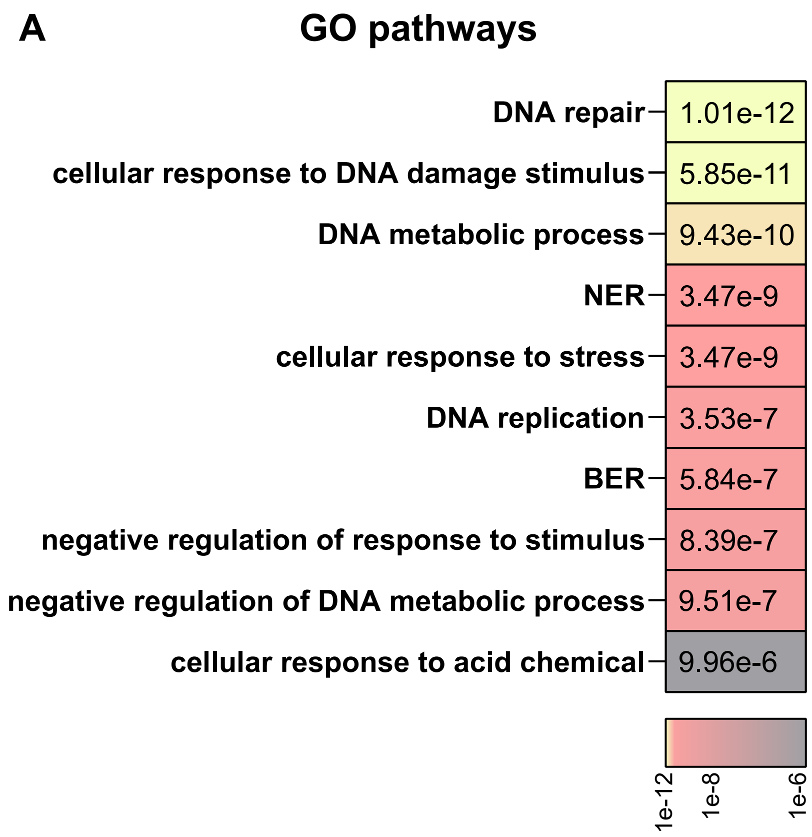


**Demidova et al., Supplementary Figure 1**

### supp fig 2

A

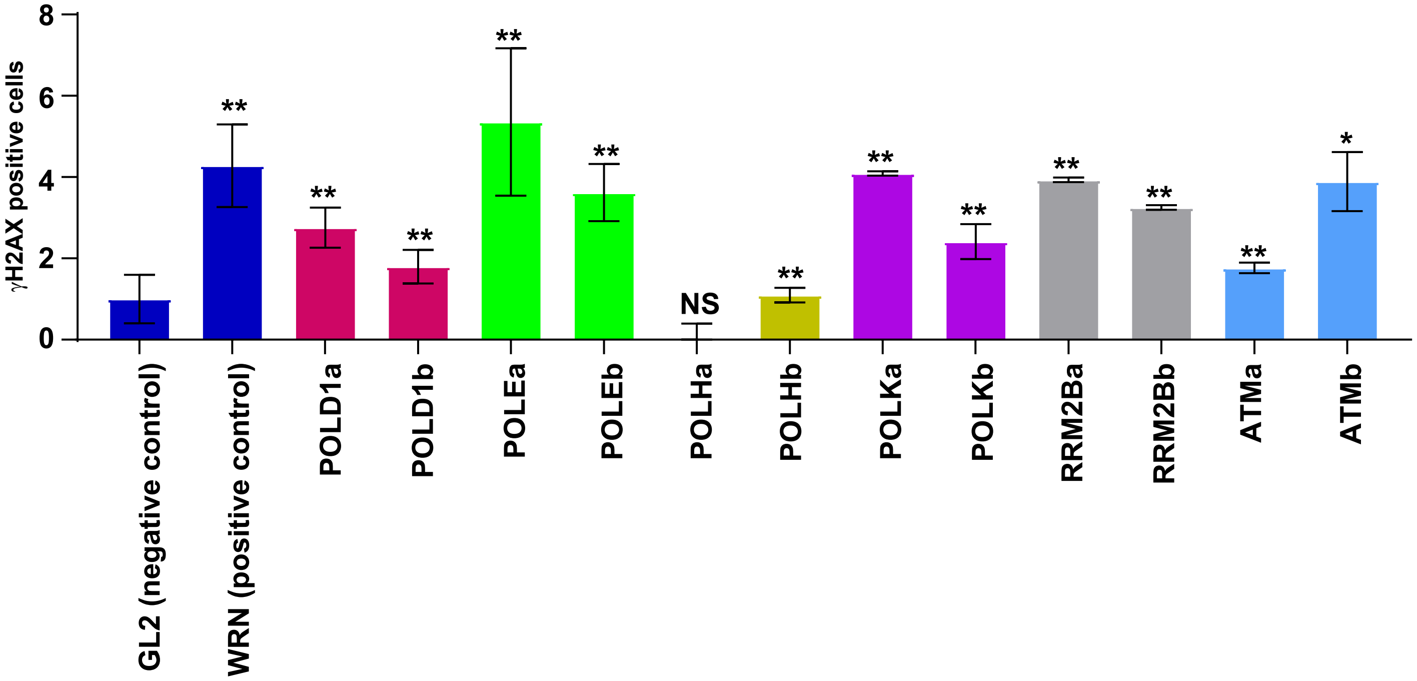


Demidova et al., Supplementary Figure 2

### supp fig 3

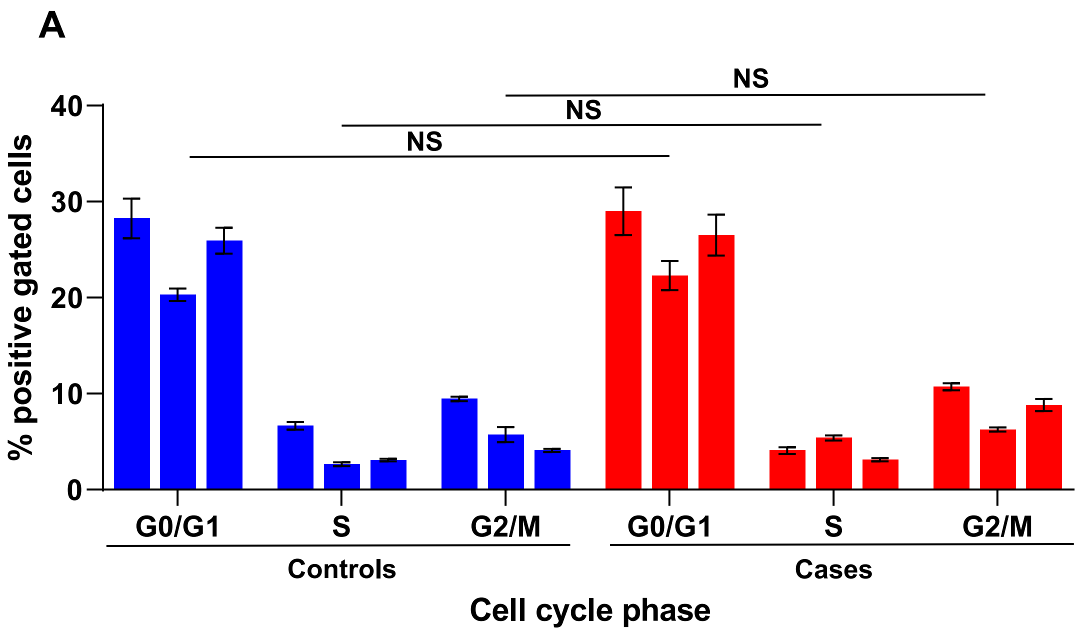


Demidova et al., Supplementary Figure 3

### supp fig 4

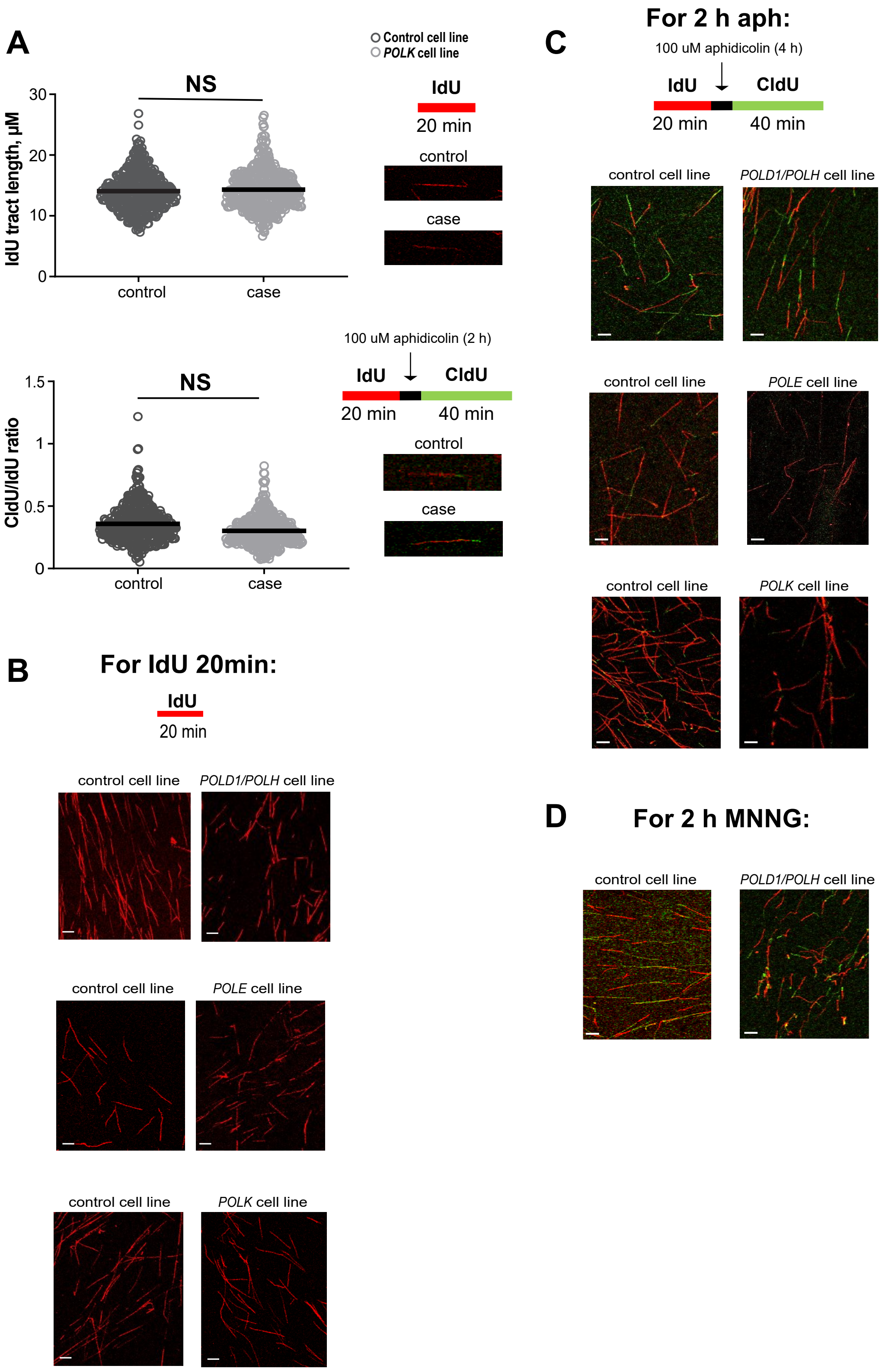


**Demidova et al., Supplementary Figure 4**

### supp fig 5

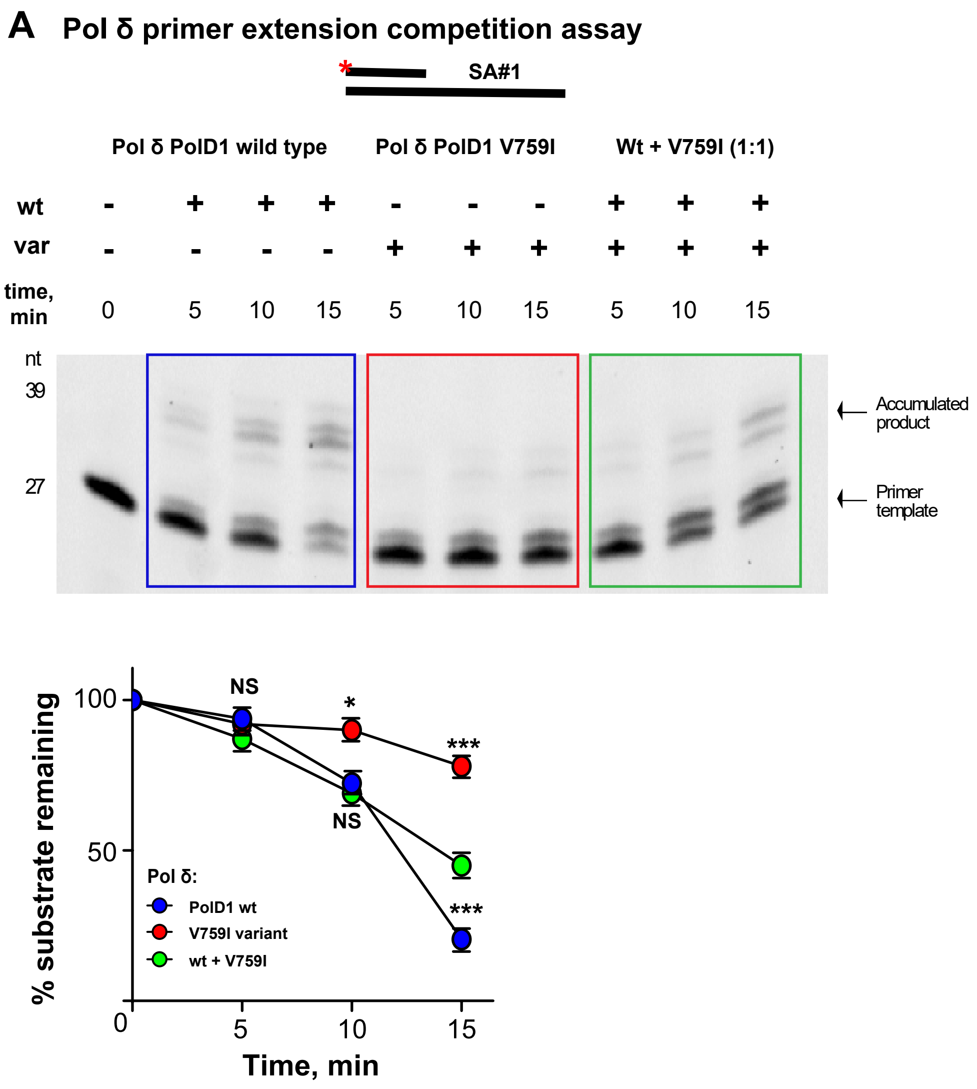


Demidova et al., Supplementary Figure 5
