## Supplementary material for "Signatures of defective DNA repair and replication in early-onset renal cancer patients referred for germline genetic testing": supp table 3

**Supplementary Table 3. Complete summary of results for the DNA polymerase variants identified in the eoRCC patients.**

| Gene | *POLD1* | *POLH* | *POLE* | *POLK* |
|---|---|---|---|---|
| **Variant** | V759I | G209V | W1624X | E29K |
| **γH2AX in patient primary PBMCs** | increased | | increased | increased |
| **γH2AX on knockdown in Caki cells** | decreased | decreased | decreased | decreased |
| **Protein expression in EBV-cell lines** | no difference | decreased | no difference | no difference |
| **Cell counts in EBV-cell lines** | no difference | | no difference | no difference |
| **CTB viability in EBV-cell lines** | increased viability | | increased viability | increased viability |
| **DNA fiber assay in EBV-cell lines** | slower replication speed, slower fork restoration | | slower replication speed, slower fork restoration | no difference in replication |
| **Cell cycle in EBV-cell lines** | no difference | | no difference | no difference |
| **Tumor sequencing (LOH)** | no LOH | no LOH | no LOH | no LOH |
| **TMB analysis** | 12.85 mut/Mb | | 14.44 mut/Mb | - |
| **Structural modeling** | may disrupt D757 residue that coordinates $Mg^{2+}$ ions in the active center of the polymerase domain | may alter stability of the α-helix in the polymerase active center | N/A (stop gain variant) | - |
| **Biochemical assays** | less robust polymerase activity, impaired function versus wild type complex | slower lesion bypass, suggestive of better processivity over wild type | Same as above | see below |
| **Biochemical assays previously reported** | - | - | - | reduced catalytic efficiency and reduced replication fidelity versus wild type(20) |

CTB – cell titer blue; DDR – DNA damage and repair; eoRCC – early-onset renal cell carcinoma; LOH – loss of heterozygosity; NS – non-significant; PBMCs – peripheral blood monocytes; TMB – tumor mutation burden
