## Supplementary material for "Signatures of defective DNA repair and replication in early-onset renal cancer patients referred for germline genetic testing": supp table 7

-in:file:s /home/andrakem/Documents/clients/Arora/ddgPolE/min_cst_0.5.AF2hPolE_0001.pdb # the PDB file of the structure on which point mutations should be made
-resfile /home/andrakem/Documents/clients/Arora/ddgPolE/mutfilePolE # the list of point mutations to consider in this run
-ddg:weight_file soft_rep_design # Use soft-repulsive weights for the initial sidechain optimization stage
-database /home/dunbracklab/Rosetta/main/database #the full path to the database is required

-ddg::iterations 25 # 50 is the recommended number of iterations
-ddg::dump_pdbs true # write out PDB files for the structures, one for the wildtype and one for the pointmutant for each iteration
-ddg::local_opt_only false # recommended: local optimization restricts the sidechain optimization to only the 8 A neighborhood of the mutation (equivalent to row 13)
-ddg::min_cst false # use distance restraints (aka constraints) during the backbone minimization phase
-constraints::cst_file /home/andrakem/Documents/clients/Arora/ddgPolE/input.cst # the set of constraints to use during minimization which should reflect distances in the original (non-pre-relaxed) structure

-in::file::fullatom # read the input PDB file as a fullatom structure
-ddg::mean true # do not report the mean energy
-ddg::min true # report the minimum energy
-ddg::sc_min_only false # do not minimize only the backbone during the backbone minimization phase
-ddg::ramp_repulsive true # perform three rounds of minimization (and not just the default 1 round) where the weight on the repulsive term is increased from 10% to 33% to 100%
